# No evidence for intervention-associated DNA methylation changes in monocytes of patients with posttraumatic stress disorder or anorexia nervosa

**DOI:** 10.1101/2020.11.11.20229567

**Authors:** Elisabeth Hummel, Magdeldin Elgizouli, Jasmin Beygo, Johanna Giuranna, Maurizio Sicorello, Elsa Leitão, Christopher Schröder, Michael Zeschnigk, Svenja Müller, Dana Öztürk, Manuel Föcker, Stephan Herpertz, Johannes Hebebrand, Dirk Moser, Henrik Kessler, Bernhard Horsthemke, Anke Hinney, Robert Kumsta

## Abstract

DNA methylation patterns can be responsive to environmental influences. This observation has sparked interest in the potential for psychological interventions to influence epigenetic processes. Recent studies have observed correlations between DNA methylation changes and therapy outcome. However, most did not control for changes in cell composition from pre- to post-therapy. This study had two aims: first, we sought to replicate therapy-associated changes in DNA methylation of commonly assessed candidate genes in isolated monocytes from 60 female patients with post-traumatic stress disorder (PTSD) using targeted deep bisulfite sequencing (DBS). Our second, exploratory goal was to identify novel genomic regions with substantial pre-to-post intervention DNA methylation changes by performing whole-genome bisulfite sequencing (WGBS) in two patients with PTSD and three patients with anorexia nervosa (AN) before and after intervention. Equivalence testing and Bayesian analyses provided evidence against physiologically meaningful intervention associated DNA methylation changes in monocytes of PTSD patients in commonly investigated target genes (*NR3C1, FKBP5, SLC6A4, OXTR)*. Furthermore, WGBS yielded only a limited set of candidate regions with suggestive evidence of differential methylation pre- to post-therapy. These differential methylation patterns did not prove replicable when investigated in the entire cohort. We conclude that there is no evidence for major, recurrent intervention-associated DNA methylation changes in the investigated genes in monocytes of patients with either PTSD or AN.

**Author Summary:** Many mental health problems have developmental origin, and epigenetic mechanisms have been proposed to explain the link between stressful or adverse experiences and subsequent health outcomes. More recently, studies have begun to examine whether psychological therapies might influence or even reverse supposedly acquired DNA methylation marks. Correlations between response to therapy and DNA methylation changes in peripheral tissue have been reported; however, these results might be confounded by differences in cell composition between time points and not reflect true DNA methylation changes. Here, we attempted to replicate previous reported results in a homogenous cell population (monocytes) and further to identify novel intervention-responsive regions in the whole genome in patients with post-traumatic stress disorder (PTSD) and anorexia nervosa (AN).

Our results showed that the improvement in symptomatology in PTSD and AN patients was not reflected in changes in DNA methylation in monocytes, neither in the previously studied candidate genes nor in the regions identified by whole-genome bisulfite sequencing. This study provides evidence against DNA methylation changes in peripheral tissue following therapy, and we suggest that previous findings are most likely explained by differences in cell composition.

## Introduction

The concept of an environmentally regulated epigenome has attracted considerable attention in the mental health field [1, 2]. Many psychopathologies have developmental origins, and epigenetic processes have been suggested as targets for the effects of psychosocial adversity particularly in early life. The epigenome is an umbrella term for a range of mechanisms involved in gene regulation, including DNA methylation, histone modifications and regulation through non-coding RNA molecules [3]. Epigenetic processes are essential for normal cellular differentiation and development. Perturbations in these processes have been linked to different pathologies [4], including mental disorders [5]. Both animal models and human studies have pointed to persistent epigenetic changes in response to various stimuli. These changes, it has been argued, might reflect or mediate the long-term effects of early risk exposure such as prenatal malnutrition on metabolic outcomes [6]. Another often quoted example of epigenetic mediation is the programming of the neuroendocrine stress response by the degree of maternal care in rodents [7].

DNA methylation, which involves methylation of cytosines predominantly in cytosine-guanine (CpG) dinucleotides, has long been thought to represent an early-established and rather stable epigenetic marker. A number of recent studies have shown, however, that DNA methylation patterns are more dynamic than previously thought, and can change in response to internal signals or environmental influences [8-12]. These observations have sparked interest in the potential for psychological and nutritional interventions (e.g. in Anorexia Nervosa) to influence these biological processes.

Several studies have been published that assessed DNA methylation before and after therapeutic intervention [see 13 for review]. Most followed a candidate-gene approach and investigated genes involved in stress regulation, such as the *Glucocorticoid Receptor* (*NR3C1*) or *FKBP Prolyl Isomerase 5* (*FKBP5*) [14-16], or genes commonly investigated in psychiatric genetics studies, such as the *Serotonin Transporter* (*SLC6A4*; [17]), *Monoamine Oxidase A* (*MAOA*; [18]), and the *Brain Derived Neurotrophic Factor* (*BDNF*; [19]). The studies conducted so far are very heterogeneous in terms of investigated disorders (post-traumatic stress disorder, PTSD; anorexia nervosa, AN; anxiety disorders, borderline personality disorder), analyzed tissue (buccal cells, whole blood, peripheral blood mononuclear cells (PBMCs), as well as type of intervention and methodology for exploring DNA methylation. A common feature of all investigations is the observation that therapy responders and non-responders showed differential DNA methylation patterns pre- to post-intervention. For instance, in combat veterans with PTSD, DNA methylation levels of *FKBP5* in PBMC decreased in responders but increased in non-responders comparing pre- to post-therapy measurements [16]. The first epigenome-wide investigation of intervention-associated DNA methylation changes in PTSD further supported this notion. Successful psychotherapeutic treatment of PTSD was associated with increases in DNA methylation of the *Zinc Finger Protein 57* gene (*ZFP57*) in whole blood, whereas DNA methylation in this region decreased during the development of PTSD, and also decreased in patients who did not respond to therapy [20]. A similar picture emerged from an epigenome-wide study of AN. Here, longer duration of illness was associated with lower DNA methylation levels in whole blood, whereas DNA methylation increased at the same positions with a concomitant increase in body mass index (BMI) across therapy [21].

Taken together, DNA methylation changes have thus been suggested as a marker or epigenetic correlate of therapy outcome. Caution is needed in the interpretation of these findings though. With exception of the two epigenome-wide association studies, only a limited number of CpGs were investigated, the observed changes were small, and the applied methods mostly had insufficient sensitivity to reliably assess such small DNA methylation differences. Furthermore, it is unclear whether differences in DNA methylation, as observed after the intervention, do not rather reflect changes in the cellular composition of the investigated tissue. DNA methylation patterns are highly cell type-specific, and each cell type within a tissue contributes to DNA methylation variation [22].

Given that the cell composition of the circulating leukocyte pool is dynamic and influenced by various external factors, including infections, menstrual cycle [23] or stress exposure [24], DNA methylation changes in blood collected at different time points from the same individual might primarily reflect differences in cell composition [25], unrelated to the effect of intervention.

To address these limitations, we set out to investigate intervention-associated changes in DNA methylation in a homogenous cell population (CD14^+^ monocytes), in a female-only PTSD patients cohort. Monocytes were chosen as previous studies have shown that among the heterogeneous leukocyte population, monocytes were the most sensitive subtype for traumatic experiences and variation of psychosocial conditions [26].

The two goals of this study were, first, to replicate findings from previous reported intervention-associated candidate genes (*NR3C1, FKBP5, SLC6A4, Oxytocin Receptor* (*OXTR)* in a female-only PTSD cohort, and, second to identify new differentially methylated regions (DMRs) by performing an exploratory whole-genome bisulfite sequencing (WGBS). For the exploratory WGBS we not only analyzed a sub-sample of the PTSD cohort but also examined a second independent female AN cohort. AN has a metabolic component and therefore we expected to find more DMRs. DNA methylation of commonly assessed candidate genes and promising targets from WGBS were analyzed using targeted deep bisulfite sequencing (DBS). Our approach thus addressed essential confounders, namely sex, cell heterogeneity, genetic variation, coverage of the genome and coverage of CpGs within candidate genes.

## Results

### Study 1 - Targeted analysis of previously investigated candidate genes

To address our first goal of replicating intervention-associated DNA methylation changes of previously assessed candidate genes, we investigated sixty female PTSD patients. Patients with PTSD had a clinically relevant reduction in PTSD symptoms, with a mean reduction of 15.6 ± 14.6 (SD) on the PTSD-Check List [PCL-5; 27]. Thirty-six patients were classified as responder according to PCL-5 (drop by 10 points from pre- to post-treatment). The responders showed a mean difference between post- and pre-treatment in PCL-5 of −24.1 ± 10.3, while the non-responders (n=21, 35%) showed a mean difference of −1.0 ± 7.8 (PCL-5 data was missing for three patients).

DBS was performed for four candidate genes for which psychotherapy effects on DNA methylation change have been previously reported: *NR3C1, FKBP5, SLC6A4* and *OXTR* (see S1 Fig for exact chromosomal locations). Linear regression analysis showed no significant association between pre-treatment mean DNA methylation of DBS targets and the severity of baseline PTSD symptoms (S3 Table). Likewise there were no statistically significant differences in mean DNA methylation between pre- and post-intervention for all investigated targets, even without correction for multiple comparisons (all *p* > .05; Table 1). Intervention-associated changes in DNA Methylation of single CpGs did not survive correction for multiple testing (S4 Table). Bayes factors favored the null hypothesis of no DNA methylation change for *NR3C1 and FKBP5*. Furthermore, DNA methylation change was not conditional on therapy response (all p > 0.16; Table 1), with all Bayes factors larger than one and Bayes factors for *SLC6A4* and *FKBP5* above the threshold of three. Moreover, we observed no statistically significant correlation between symptom changes and DNA methylation changes (all p > 0.15; Table 2 and S2 Fig, upper row), with all Bayes factors larger than one and Bayes factors for *SLC6A4* and *FKBP5* above three. Distributions of DNA methylation changes are shown in Fig 1, upper row by responder status. The largest mean differences between time points was found for *OXTR* (−0.26% ± 1.21%), and the smallest was −0.019% ± 0.19% for *NR3C1*. Mean DNA methylation levels pre- and post-intervention averaged across the analyzed genomic regions are shown in S3 Fig.

**Table 1.**
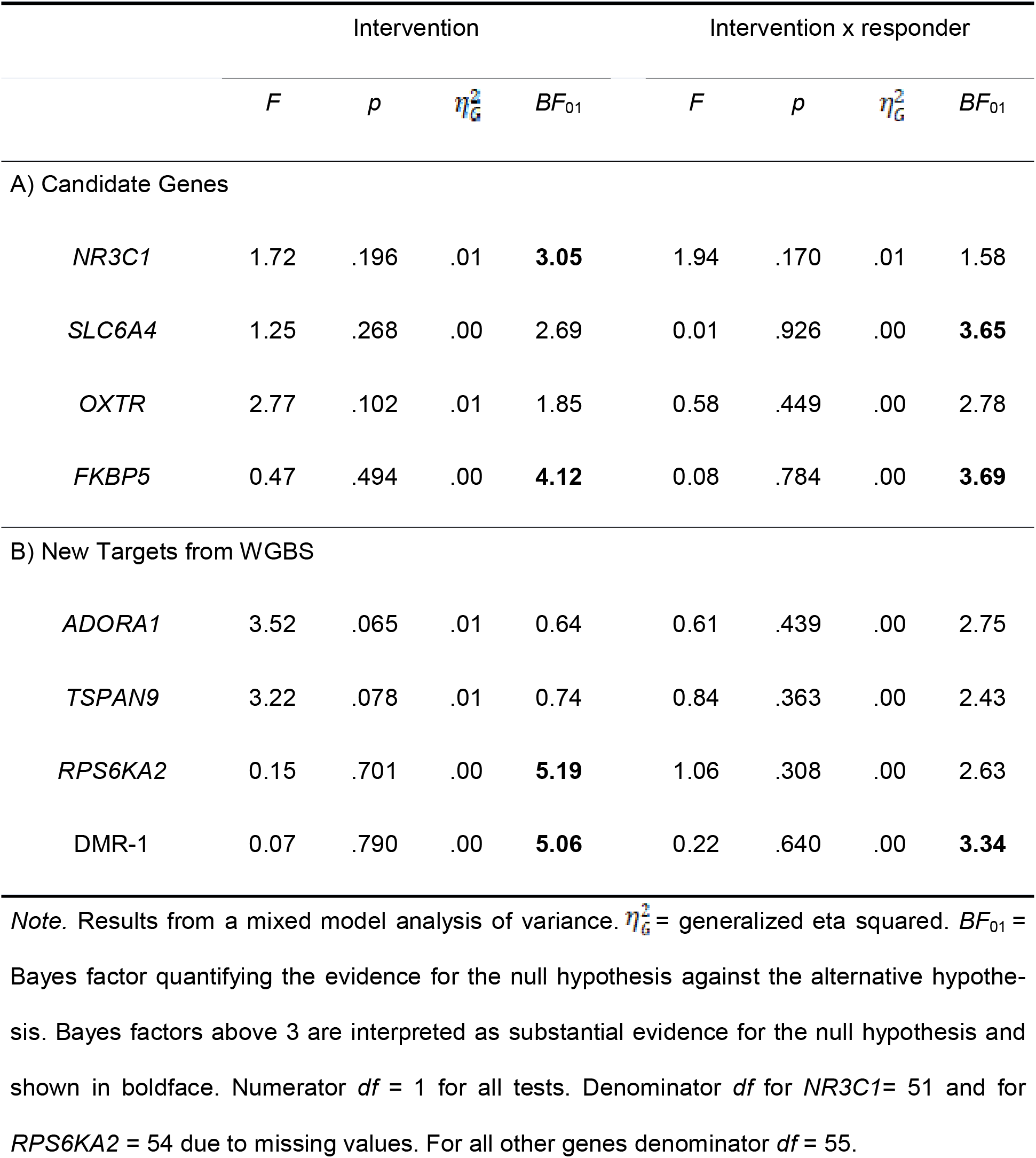
**Effect of *intervention* and the effect of *intervention* by *responder* status interaction on DNA methylation in the PTSD cohort**

**Table 2.** **Correlations between change in DNA methylation and change in symptom scores (PTSD cohort)**.

| Gene | *r* | 95% CI | *p* | *BF*<sub>01</sub> |
| --- | --- | --- | --- | --- |
| A) Candidate Genes |  |  |  |  |
| *NR3C1* | -.17 | [-.42, .11] | .235 | 1.70 |
| *SLC6A4* | -.02 | [-.28, .24] | .874 | **3.30** |
| *OXTR* | .11 | [-.16, .36] | .425 | 2.50 |
| *FKBP5* | -.04 | [-.30, .22] | .778 | **3.22** |
| B) New Targets from WGBS |  |  |  |  |
| *ADORA1* | -.19 | [-.43, .07] | .155 | 1.32 |
| *TSPAN9* | .16 | [-.11, .40] | .242 | 1.78 |
| *RPS6KA2* | -.12 | [-.37, .14] | .365 | 2.28 |
| DMR-1 | -.10 | [-.35, .17] | .473 | 2.64 |

*Note.* *r* = pearson correlation. *BF*<sub>01</sub> = Bayes factor of For *NR3C1*, *n* = 53. For *RPS6KA2*, *n* = 56. For all remaining genes, *n* = 57. *BF*<sub>01</sub> = Bayes factor quantifying the evidence for the null hypothesis against the alternative hypothesis. Bayes factors above 3 are interpreted as substantial evidence for the null hypothesis and shown in boldface.

**Fig 1.**
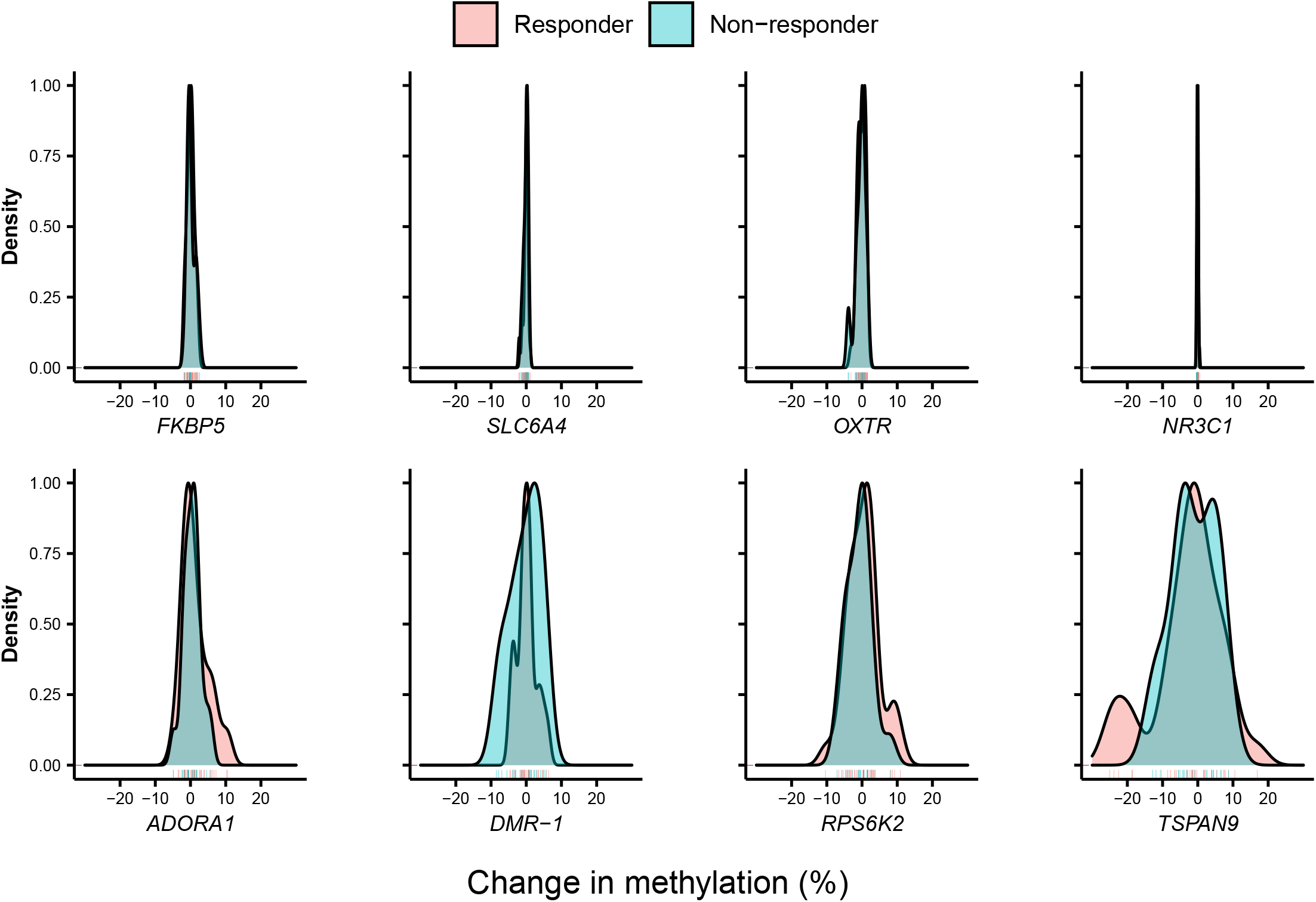
**Density plots for the distribution of DNA methylation change (post minus pre-treatment) by therapy response and gene.**

Complementary to the Bayes factor approach, equivalence test help infer whether true population effect sizes are smaller than a pre-defined smallest effect size of interest. Effect sized for all candidate genes were significantly smaller than 1% DNA methylation change, even in the therapy-responder group and after Bonferroni correction (all *p*_*corrected*_ < .001). The 95% confidence intervals of difference scores in the responder group did not include methylation changes larger than ≈|0.5%|: Δ_FKBP5_ = 0.06% [-0.28, 0.40], Δ_SLC6A4_ = −0.11% [-0.33, 0.11], Δ_OXTR_ = −0.16% [-0.52, 0.21], Δ_NR3C1_ = 0.00% [-0.06, 0.07].

We conclude that true effect sizes of all classic candidate genes are smaller than 1% DNA methylation change.

### Study 2 − Explorative whole-genome bisulfite sequencing

In addition to our goal of conceptually replicating previously reported results in candidate genes, we aimed to explore whether we could identify genomic regions with large pre-post intervention changes in DNA methylation in a limited sample, which could then be validated in the larger sample. Besides the PTSD patients, we included AN patients for explorative WGBS. Due to the strong metabolic component during AN therapy, we expected a higher number of DMRs in the AN patients. Following recommendations by Ziller et al. [28], we sequenced at least two biological replicates at >5x coverage. Pre- and post-intervention monocyte DNA from two patients with PTSD and additionally from three AN patients was subjected to WGBS. The patients for the WGBS were selected based on their responsiveness to the therapy, absence of co-morbidity, and non-smoking status.

There was a slight difference in the genome-wide mean methylation levels between the PTSD and AN samples (0.72 vs. 0.75; Mann-Whitney U test; S5 Table). DNA methylation did not differ between pre- and post-intervention samples of the same individuals (p-value = 1; Wilcoxon signed rank test).

A principal component analysis (PCA) of the PTSD and AN methylomes did not group samples according to pre- or post-intervention state (S4 Fig). The same was observed in a cluster analysis of the 1,000 most variable CpGs for the PTSD and AN datasets, in which separate branches are observed for each individual (Fig 2). We observed large differences between individuals, most likely reflecting the different genetic backgrounds, but no recurrent differences between the pre- and post-intervention states.

**Fig 2.**
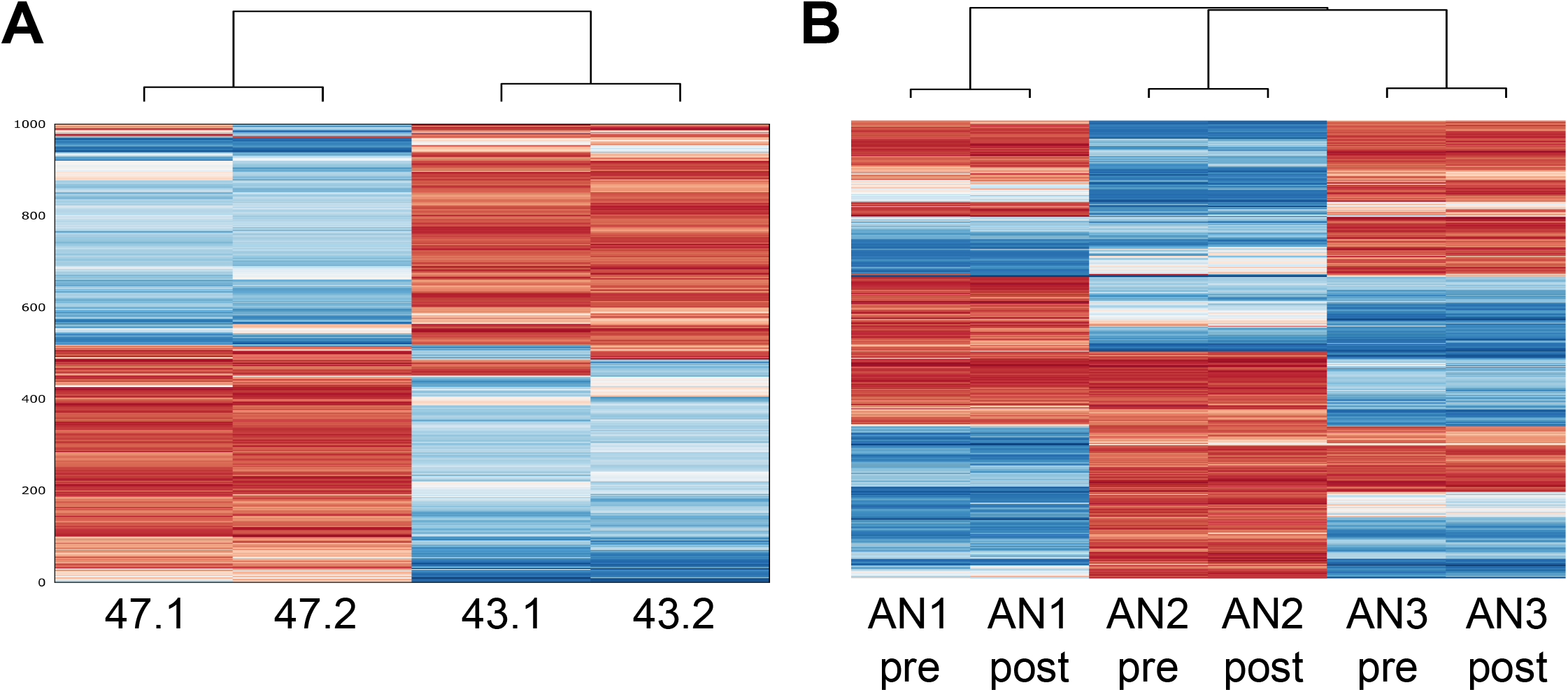
Cluster analysis of 1,000 most variable CpGs. A) The PTSD monocyte methylomes were obtained from two individuals (47 and 43), pre- (.1) and post-intervention (.2). B) The AN monocyte methylomes were obtained pre- and post-treatment from three individuals (AN1, AN2 and AN3). CpG SNPs were excluded from the analysis. The difference between individuals is much greater than between pre- and post-intervention.

To identify DMRs between pre- and post-treatment samples from each cohort, we used two different bioinformatic tools (camel and metilene; [29, 30]). Owing to lower coverage in the PTSD samples compared to the AN samples, we set the threshold to a minimum coverage of five reads and ten reads, respectively. Defining a camel DMR as a region with at least four CpGs and a minimum methylation difference of 0.3, we detected 33 DMRs in the PTSD patients (S6 Table). Using a filter of q < 0.05 for metilene DMRs, we detected four DMRs (S7 Table).

Using the same DMR filters, camel identified no DMRs in the AN data set, indicating that there are no major recurrent methylation changes. Since the samples of this cohort were sequenced at ahigher depth, we decided to lower the minimum methylation difference between groups to 0.2 while at the same time increasing stringency by setting the minimum number of CpGs to 10. Using these filters, we identified 35 DMRs with camel (S8 Table). At q<0.05, metilene reported 10 DMRs, with seven DMRs being detected by both tools (S9 Table). There is one DMR common to the PTSD and AN datasets, which overlaps the binding sites for CTCF and other transcription factors within intron 1 of *EXD3*, a gene encoding an exoribonuclease required for 3’-end trimming of AGO1-bound miRNAs.

### Validation of WGBS-nominated targets in the PTSD sample

DNA methylation pattern of four targets that emerged as potentially responsive to intervention from WGBS data were validated in the entire sample (n=60) using DBS. The following gene regions were analyzed: DMR-4 within *Adenosine A1 Receptor* (*ADORA1)*, DMR-21 within *Tetraspanin 9 (TSPAN9)*, DMR-10 within *Ribosomal Protein S6 Kinase A2 (RPS6KA2)*, and intergenic DMR-1 (S6 Table). Chromosomal locations of the targets are shown in S1 Fig.

The effect sizes of *ADORA1, RPS6KA2*, and DMR-1 were all significantly smaller than 5% DNA methylation change in both the whole cohort and the responder cohort (all *p*_*corrected*_ < .001). The effect of *TSPAN9* was only significantly different from 5% in the whole cohort on an uncorrected *p* = .043, which did not survive Bonferroni correction. There were no statistically significant differences in DNA methylation between pre- and post-intervention for the WGBS-nominated targets. However, the possibility of a true effect size within the range of 5% DNA methylation difference at the *TSPAN9* between pre- and post-intervention samples cannot be ruled out. Bayes factors favored the null hypothesis of no methylation change for *RPS6KA2* and DMR-1. Similar to candidate genes, DNA methylation change was not conditional on therapy response (all p > 0.05; Table 1), with all Bayes factors larger than one and Bayes factors for DMR-1 above the threshold of three. There were no statistically significant correlations between symptom changes and DNA methylation changes (all p > 0.05; Table 2 and S2 Fig, lower row), with all Bayes factors larger than one. Distributions of DNA methylation changes are shown in Fig 1, lower row by responder status. The largest mean differences between time points was found for *TSPAN9* (−2.78% ± 9.78%), and the smallest was −0.12% ± 3.35% for DMR-1. Mean DNA methylation levels pre- and post-intervention averaged across the analyzed genomic regions are shown in S3 Fig. DNA methylation levels for single CpGs are shown in S4 Table.

Equivalence tests showed that the effect sizes of *ADORA1, RPS6KA2*, and DMR-1 were all significantly smaller than 5% DNA methylation change in both the whole cohort and the responder cohort (all *p*_*corrected*_ < .001). The effect of *TSPAN9* was only significantly different from 5% in the whole cohort on an uncorrected *p* = .043, which did not survive Bonferroni correction. Hence, although we cannot rule out that there is a true effect size as large as 5% DNA methylation change for the *TSPAN9* locus, we can conclude with relative certainty that true effect sizes of the WBGS nominated genes are smaller than 5%.

### Validation of WGBS-nominated targets in the AN sample

In the AN cohort, DBS was performed for validation of targets that emerged from WGBS (methylation difference ≥ 0.2) in all three individuals as potentially responsive to weight gain in eight regions (four detected by both DMR-calling tools and four detected by camel: DMR-1 within *Galactosidase Beta 1 Like* (*GLB1L*), intergenic DMR-3, DMR-7 within *Family With Sequence Similarity 50 Member B* (*FAM50B*), DMR- 11 within *Mesoderm Specific Transcript* (*MEST*), DMR-13 within *ER Lipid Raft Associated 2* (*ERLIN2*), DMR-16 within *Exonuclease 3’-5’ Domain Containing 3* (*EXD3*), DMR-22 within *SNRPN Upstream Reading Frame/ Small Nuclear Ribonucleoprotein Polypeptide N* (*SNURF*/*SNRPN*) and DMR-28 within *Heat Shock Protein Family A (Hsp70) Member 12B* (*HSPA12B*) (for DMR numbering see S8 Table). All CpGs contained in the DMRs were included in the amplicons. Chromosomal locations are shown in S2 Table.

Since for this cohort we had used a lower minimum methylation difference between groups (0.2), we first attempted technical confirmation using DBS in the same three individuals (pre- and post-treatment; same DNA preparation as for WGBS). As can be seen in S5 Fig, the methylation differences identified by WBGS could not be confirmed in any of these regions. Therefore, the four independent samples were not further analyzed.

## Discussion

Epigenetic processes have been proposed as a potential mechanism mediating the link between exposure to trauma or adversity and mental health problems. Expectedly, there is great interest in the question whether psychological interventions translate into a modification of disorder-related epigenetic signatures. The first few studies addressing this question have found divergent patterns of DNA methylation changes between patients who responded to therapy and those who did not. However, results of these pilot studies remain inconclusive, because differences in genetic background and changes in cell composition over time, which are the main confounders in epigenetic research, were not taken into account.

In our study, we avoided these confounders by using isogenic pre- and post-treatment samples and a homogenous cell population (monocytes). Furthermore, we used DBS for targeted high resolution methylation analysis. With this approach, we could not identify any evidence of intervention-associated changes in DNA methylation. We investigated four candidate genes that were previously shown to be sensitive to variation in the social environment and/or seemed responsive to intervention in a sample of female PTSD patients. The prime candidate for epigenetic research in humans has been *NR3C1*, since DNA methylation of the promoter of alternative Exon 1_7_ (1F in humans) had been shown to vary as a function of maternal care in the rodent model [7]. We found no change between pre- and post-intervention in PTSD patients and no differences between therapy responders and non-responders. We also investigated DNA methylation of *SLC6A4*, the most widely studied candidate gene in psychiatry. One previous study found small increases in *SLC6A4* methylation in therapy responders in one of four assessed CpG sites [17]. Here, we investigated 82 CpGs sites across the entire CpG island in the gene’s promoter region and found no evidence for change over time - neither overall nor when stratified by responders and non-responders. This was also true for *FKBP5*, where we focused on 5 CpGs in intron 7, previously associated with PTSD risk following early life adversity [31], and the *OXTR* gene, where we investigated 16 CpGs in a putative enhancer region - characterized by H3K27 acetylation peaks - in intron 3 of the gene. In sum, we did not observe intervention-associated changes in DNA methylation of commonly investigated candidate genes when assessed in a homogenous cell population.

In small samples with limited statistical power, non-significant results do not provide evidence for the absence of meaningful effects. We thus further conducted equivalence tests to ascertain whether observed effect sizes are significantly smaller than a minimal effect size of interest. These revealed that DNA methylation changes of the four candidate genes were significantly below physiologically meaningful levels [32, 33]. Notably, this does not preclude that miniscule changes on these genes might contribute to a meaningful epigenome-wide poly-epigenetic score, but such studies necessitate much larger samples and another theoretical perspective on the importance of single candidate genes than the studies whose effects we aimed to replicate.

Candidate gene studies can only provide a very limited view on potential dynamics of the epigenome. True epigenome-wide studies covering the ∼28 million CpG sites across the human genome are still prohibitive in large cohorts because of the associated costs. Here, we used an explorative strategy to identify genomic regions whose DNA methylation pattern might potentially be responsive to intervention, by using WGBS in two PTSD and three AN patients before and after therapy. We identified a small number of pre-to-post intervention DMRs (36 for PTSD and 38 for AN). The validation of a subset of the identified PTSD DMRs in the remaining 58 patients could not confirm the potential differential methylation patterns found by WGBS. In addition, AN DMRs identified after reducing the stringency in DMR calling (by lowering to 0.2 the minimum methylation difference allowed) could not be validated by DBS in the three samples subjected to WGBS.

It is remarkable that the change of physiological conditions for monocytes in AN due to adaption to prolonged starvation is not associated with significant changes in DNA methylation pattern. This suggests that cell type-specific methylation patterns are highly stable. This stability can probably be attributed to stem or progenitor cells, because monocytes have a very short half-life in blood. Pre- and post-treatment samples are not drawn from the same pool of monocytes, but are derived from the same stem or progenitor cells. We previously showed differences in metabolic profiles in patients with AN compared to controls and in patients with AN between acute stage of starvation and after short term weight recovery. A substantial number of metabolite levels did not return to normal after short term recovery [34]. It might be assumed that weight gain could exert an effect on the methylome at a later time point.

It is important to reflect on the question whether it is plausible to expect changes of DNA methylation patterns in peripheral surrogate tissues that are associated with changes in thoughts, feelings or somatic alterations in a meaningful way. Three scenarios have been put forward previously: first, DNA methylation dynamics observed in blood or buccal epithelial cells reflect the processes occurring in neuronal cells [18]. However, as buccal epithelium or leukocytes do not have a biologically realistic link to cellular processes occurring in neurons, these transient and activity-dependent alterations of chromatin and the specific patterns of DNA methylation in neurons are most likely not reflected in peripheral tissue.

The second scenario is that changes in peripheral DNA methylation patterns are brought about by intervention-associated psychophysiological changes that parallel changes in thoughts and feelings. Such cross talk between the central nervous system and peripheral organs is arguably mediated through regulation of the autonomic nervous system and the hypothalamic–pituitary–adrenal axis, activating the two main stress effectors noradrenaline and cortisol. These mediate their effects by engaging cellular receptor systems, which ultimately regulate specific gene expression responses. A testable model therefore suggests that changes in DNA methylation are brought about by alterations of transcriptional activity associated with changes in upstream signaling of stress mediators such as cortisol or noradrenalin [35].

The third scenario, which is in our opinion the most plausible explanation for many previous findings, is that changes in DNA methylation levels merely reflect changes in cell composition, which might be stochastic in nature, reflect differences in health status independent of disorder of interest, or reflect intervention-associated physiological changes. As such, change in the composition of the leukocyte pool might be a marker of therapy response [36].

The following limitations deserve a mention. The costs associated with WGBS limited the number of patients to 2-3 for each cohort, so that only large pre-to-post differences could be reliably identified. Power estimates show that we had 80% power to detect pre-to-post-treatment DNA methylation differences of 30%. Furthermore, only one cell type was investigated. Monocytes have been shown as the most sensitive subtype for social conditions and traumatic experiences, at least in terms of transcriptional changes [26, 37-39]. However, any effects in other cell types (e.g., T cells, B cells, NK cells, etc.), or changes in the relative prevalence of cells in the circulating leukocyte pool are missed in this analysis.

In conclusion, our results provide no evidence in support of intervention-associated changes in DNA methylation in monocytes in PTSD or AN, and equivalence tests and Bayesian statistics provided evidence against even subtle DNA methylation changes associated with therapy outcome. We argue that the previously reported changes in DNA methylation following therapy are most likely explained by changes in cell composition rather than by cellular reprogramming of DNA methylation. In itself, any shifts in cellular composition might reflect intervention-associated physiological changes and could therefore be used as biomarkers, but markers should not be confused with mechanisms.

## Methods

### Sample Characteristics, clinical judgement and treatment

#### PTSD cohort

We investigated sixty female in-patients of European descent seeking treatment for PTSD at the Department of Psychosomatic Medicine and Psychotherapy, LWL-University Hospital, Ruhr-University Bochum. The patients were between 20 and 60 years old (mean age: 40.0 ± 11.9

(SD) years), and the mean treatment duration was 6.5 ± 1.4 (SD) weeks. Inclusion criteria were PTSD diagnosis and female sex. The PTSD study was approved by the ethics committee of the Faculty of Psychology, Ruhr University Bochum (Nr. 155), and patients gave written informed consent (see S1 Table for more details).

All patients with PTSD were diagnosed with ICD-10 (F43.1; International Classification of Diseases, 10th revision, WHO, 1993) prior to in-patient admission via structured clinical interviews in the outpatient department of the hospital. Furthermore, symptoms of PTSD were recorded before and after in-patient treatment with the PCL-5.

Participants of the PTSD cohort received standard in-patient PTSD treatment. This involved one session each week of individual cognitive-behavioral therapy, three sessions of trauma group therapy, two sessions of trauma stabilization group therapy, one session of a “skills group”, two sessions of kinesitherapy, two sessions of art therapy, physiotherapy, clinical rounds, and daily short sessions with a nurse. During individual therapy sessions patients received different trauma exposure methods. Overview of the study design is shown in Fig 3.

**Fig 3.**
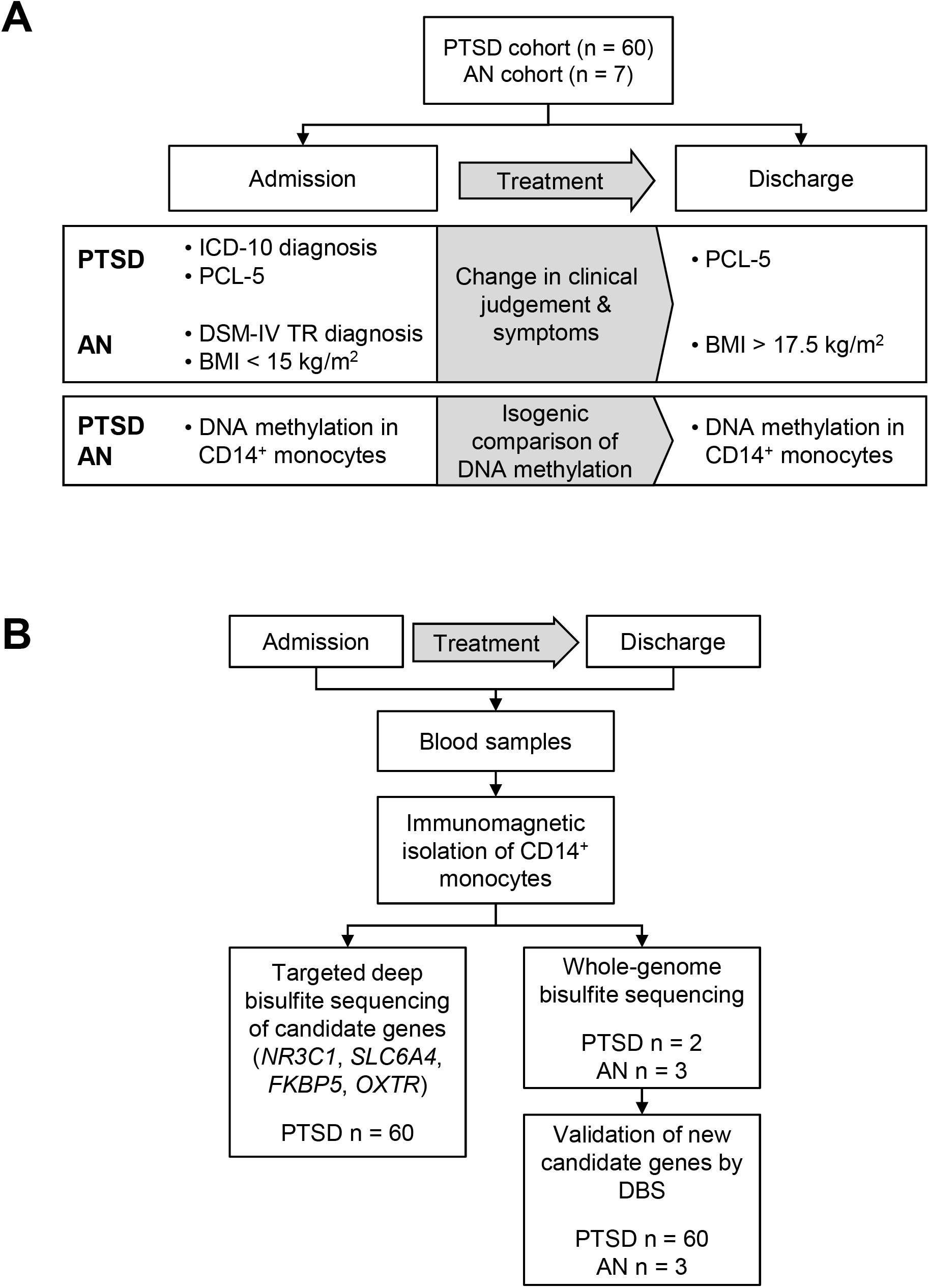
Overview of the study design.

#### AN cohort

The second cohort comprised seven female adolescent patients with AN, receiving in-patients treatment at the University Hospital Essen, University of Duisburg-Essen. The median age was 16 (with first and third quartile: Q1, Q3; 15, 17) years, median height 162.5 (160.2, 167.7) cm, median weight at admission 40.2 (37.6, 46.7) kg, median BMI at admission 15.2 (14.8, 16.1) kg/m^2^. Length of treatment was 94 (69, 99) days (see S1 Table 1 for sample characteristics). Inclusion criteria for the AN group were a diagnosis of AN, female sex, German ancestry and age from 12 to 18 years. The AN study was approved by the ethics committee of the Faculty of Medicine, University Hospital Essen (Nr. 06-3212 amendment of November 2016). Patients and their parents gave written informed consent. Patients with AN were examined at the Department of Child and Adolescent Psychiatry, Psychosomatics and Psychotherapy, University Hospital Essen, University of Duisburg-Essen according to Diagnostic and Statistical Manual of Mental Disorders, fourth edition, text revision (DSM-IV-TR) criteria [40]. AN diagnosis was confirmed via clinical examination and a semi structured interview (Kiddie Schedule for Affective Disorders and Schizophrenia (K-SADS [41]). Adolescent patients with AN participated in a multimodal multidisciplinary treatment program based on dialectical behavioral therapy for adolescents that included weight restoration, individual nutritional counseling, individual therapy (twice a week), skills-groups and individual skill training (twice a week), family sessions (fortnightly), and a group psychoeducation program for parents (monthly). During weight restoration, patients received nutritional counselling and meal support training. Therapy goals were a weight gain of at least 500g per week and a minimum target weight in the range of the 10-20th age-adjusted BMI percentile.

Patients with AN had a median BMI at admission of 15.2 (14.8, 16.1) kg/m^2^ and median BMI at end of treatment was 17.5 (17.5, 17.8) kg/m^2^. During the treatment period (i.e. hospital stay) of a median of 94 (69, 99) days, all seven patients gained considerable weight (median weight gain 5.8 (5.6, 6.7) kg; median BMI gain 2.2 (2.0, 2.4) kg/m^2^).

### Sample preparation and targeted deep bisulfite sequencing

Nine ml blood was drawn in the morning between 7-9 am (S-Monovette 9 ml K3E, Sarstedt). Monocytes were immunomagnetically purified from whole blood with the MACS System (Miltenyi Biotec, Bergisch Gladbach, Germany), shock frozen and stored at −80°C. DNA was isolated with the AllPrep RNA/DNA Mini Kit (Qiagen, Hilden, Germany). For DBS, DNA was bisulfite-modified with the EZ DNA Methylation Gold Kit (Zymo Research, Orange, CA, USA) and processed as described elsewhere [42, 43] using primers as indicated in S2 Table.

### Whole-genome bisulfite sequencing

For PTSD samples, WGBS libraries were prepared as previously described [44]. For the AN samples, 5 µg DNA at 50 ng/µl in water was used for library preparation and NGS sequencing on a HiSeq X platform (DKFZ Genomics & Proteomics Core Facility (GPCF) with 1 sample per lane. Read data from both datasets were processed as described previously [45, 46]. As a reference genome, we used hg19. For calling DMRs, we used camel [30] and metilene [29].

### Statistical Analyses

Repeated measures analyses of variance (ANOVA) were performed to assess changes in DNA methylation. Therapy response was included as an additional between-subject factor to check for therapy outcome-dependent changes in DNA methylation. Moreover, Pearson correlations were computed between percent DNA methylation change and PCL-5 symptom change to test a continuous measure of therapy response. In addition to changes over time, linear regression analysis was performed with pre-treatment mean DNA methylation of DBS targets and severity of baseline PTSD symptoms.

Our sample size of *N* = 60 was based on the average sample size of previous studies in therapy-contingent DNA methylation changes in mental disorders (*M* = 69, range = 16-115). Due to the heterogeneity of reported effect sizes, statistical tests, designs, and populations, a principled effect size estimation based on previous studies was not suitable in our case. We thus performed a sensitivity analysis with the software *gpower [47]*, which showed acceptable statistical power (β = .80) to detect small changes from pre- to post-treatment at η^2^ = .03, assuming a correlation between pre- and post-intervention methylation of *r* = .50. For comparisons between responders and non-responders or correlations between symptom change and methylation change, moderate effect sizes at η^2^ = .09 can be detected with sufficient statistical power, which is still in the range of previously reported effect sizes [13].

Generally, non-significant results do not provide evidence for the null hypothesis (i.e. absence of effect), especially when the statistical power is limited. Therefore, p-values were supplemented with Bayes factors to quantify the evidence for the null hypothesis using the BayesFactor package (v0.9.12-4.2) in R (3.6.1) and non-informative default priors [48, 49].

Complementary to the Bayesian approach, we conducted equivalence tests to assess whether effect sizes in DNA methylation change are significantly smaller than the smallest biologically meaningful effect size. It has been argued that DNA methylation below 5% should be interpreted with extreme caution [32, 33]. We used the two one-tailed t-test procedure [50] to check whether empirical effect sizes for methylation change are smaller than 5% or even a more conservative 1%. For a detailed account of statistical methods please see S2 Methods.

## Data Availability

The WGBS datasets generated and analyzed during the current study are available in
the European Nucleotide Archive (ENA) under the accession number PRJEB38906.
DBS data and analysis script to reproduce the statistical analyses can be found on the
open science framework:
https://osf.io/eagjx/?view_only=9737755805f045edaa1d757cc3e9ba84

## Acknowledgement

We thank the Cologne Center for Genomics (CCG) and the DKFZ Genomics & Proteomics Core Facility (GPCF) for performing WGBS.

## Supporting Information

**S1 Methods. Cell homogeneity**.

**S2 Methods. Statistical analysis**.

**S1 Table. Sample Characteristics**.

**S2 Table. Primer Sequences**.

**S3 Table. Linear regression with pre-treatment mean DNA methylation and severity of baseline PTSD symptoms**.

**S4 Table. Mean DNA methylation levels at single CpG sites**.

**S5 Table. WGBS descriptive information**.

**S6 Table. DMRs in the PTSD cohort identified with camel**.

**S7 Table. DMRs in the PTSD cohort identified with metilene**.

**S8 Table. DMRs in the AN cohort identified with camel**.

**S9 Table. DMRs in the AN cohort identified with metilene**.

**S1 Fig. Chromosomal location of analyzed regions of the PTSD cohort**.

**S2 Fig. Correlations between symptom change and DNA methylation change in the PTSD cohort**.

**S3 Fig. Mean DNA Methylation of the PTSD cohort**.

**S4 Fig. Principal component analysis**.

**S5 Fig. DNA methylation differences of the AN cohort**.

