## Supplementary material for "No evidence for intervention-associated DNA methylation changes in monocytes of patients with posttraumatic stress disorder or anorexia nervosa": Supporting Information_2021-02-24_V2.pdf

### 1. Supporting Methods

#### 1.1 S1 Methods. Cell homogeneity

The homogeneity of the cells in the PTSD cohort was determined with the BD FACSCanto TM II Flow Cytometer (BD Biosciences, San Jose, CA, USA), and showed high purity (98.3 % ± 2.4 % (SD).

#### 1.2 S2 Methods. Statistical analysis

In the PTSD cohort, repeated measures analyses of variance (ANOVA) were performed to assess changes in DNA methylation from pre- to post intervention in selected candidate genes as well as WGBS nominated novel targets. Therapy response was included as an additional between-subject factor to check for therapy outcome-dependent changes in DNA methylation. Moreover, Pearson correlations were computed between percent DNA methylation change and PCL-5 symptom change to present a continuous measure of therapy response. P-values derived from both approaches, categorical and continuous, were supplemented with Bayes factors to quantify the evidence for the null hypothesis using the BayesFactor package (v0.9.12-4.2) in R (3.6.1) and non-informative default priors. The Bayes factor divides the likelihood of the data given a model without the effect of interest by the likelihood of the data given a model including the effect of interest. If the data is more likely under the null hypothesis, the Bayes factor becomes larger than 1. Classically, Bayes factors above 3 are interpreted to represent substantial evidence in factor of the null hypothesis, although this convention should be viewed as a rough guideline instead of a definitive cutoff.

Complementary to the Bayesian approach, we conducted equivalence tests to assess whether effect sizes in DNA methylation change are significantly smaller than the smallest biologically meaningful effect size. It has been argued that DNA methylation below 5% should be interpreted with extreme caution. We used the two one-tailed t-test procedure to check whether empirical effect sizes for methylation change are smaller than 5% or even a more conservative 1%. These tests were conducted for both the whole cohort and the subgroup of therapy-responders. This approach is useful here, as the Bayes factor approach only evaluates the evidence against the existence of a standardized effect size (e.g. Cohens d), which represent a ratio between signal and noise. If both signal and noise are small, i.e. when methylation is relatively stable over time, equivalence tests against an unstandardized theoretically meaningful effect size (i.e. percent methylation change) can be more powerful.

### 2. Supporting Tables

#### 2.1 S1 Table. Sample Characteristics

| **A) PTBS cohort** | **N** | **Min** | **Max** | **Mean** | **SD** |
|---|---|---|---|---|---|
| **Age [years]** | 60 | 20 | 60 | 40 | 11.86 |
| **BMI [kg/m²]** | 60 | 17.8 | 45.5 | 30.1 | 7.26 |
| **treatment duration [days]** | 60 | 20 | 69 | 45.35 | 9.87 |
| **PTSD symptoms pre (PCL-5)** | 58 | 22 | 77 | 55.87 | 11.53 |
| **PTSD symptoms post (PCL-5)** | 59 | 5 | 80 | 40.37 | 16.92 |
| **Leukocytes pre [cells/nl]** | 59 | 3.5 | 16.3 | 7.8 | 2.52 |
| **Leukocytes post [cells/nl]** | 54 | 3.7 | 13.1 | 7.3 | 2.03 |
| **Platelets pre [cells/nl]** | 59 | 125 | 524 | 282.85 | 79.72 |
| **Platelets post [cells/nl]** | 57 | 172 | 469 | 284.37 | 76.43 |
| | | **Yes** | **No** | | |
| **Smoking** | 59 | 33 | 26 | | |
| **psychotropic medication (pre)** | 57 | 48 | 9 | | |
| **psychotropic medication (post)** | 60 | 56 | 4 | | |
| **Diagnosis of PTSD** | 60 | 60 | 0 | | |
| **Diagnosis of depression** | 60 | 57 | 3 | | |
| **other psychological comorbidities** | 60 | 31 | 29 | | |
| **somatic comorbidities** | 60 | 26 | 34 | | |
| **B) AN Cohort** | **N** | **First quartile** | **Third quartile** | **Median** | |
| **Age** | 3 | 15 | 17 | 16 | |
| **Height at admission** | 3 | 160.2 cm | 167.7 cm | 162.3 cm | |
| **Weight at admission** | 3 | 37.6 kg | 46.7 kg | 40.2 kg | |
| **BMI at admission** | 3 | 14.8 kg/m² | 16.1 kg/m² | 15.2 kg/m² | |
| **treatment duration (days)** | 3 | 69 days | 99 days | 94 days | |
| **Age** | 3 | 15.6 years | 17.3 years | 16.4 years | |
| **Height after treatment** | 3 | 160.2 cm | 167.7 cm | 162.5 cm | |
| **Weight after treatment** | 3 | 44.0 kg | 52.6 kg | 46.0 kg | |
| **BMI after treatment** | 3 | 17.5 kg/m² | 17.8 kg/m² | 17.5 kg/m² | |

#### 2.2 S2 Table. Primer Sequences

All genome positions given correspond to hg19/GRCh37.

PCRs in the AN cohort were performed as described in Leitão et al. 2018. PCRs in the PTSD cohort are described below.

1.PCR: The first round PCR reaction contained 1 µl of bisulfite-converted DNA. 0.125 µM primers and 5.5 µl GoTaq® G2 Hot Start Master Mixes (Promega. Fitchburg. WI. USA) in a total volume of 11 µl. The standard amplification protocol included an initial denaturation step for 2 min at 95 °C. followed by 50 cycles of melting at 94 °C for 30 s. annealing at 54.7-61.5 °C for 45 s and extension at 72 °C for 45 s. Followed by an additional extension at 72 °C for 10 min at the end of the 50 cycles. All PCRs included nontemplate controls. The primer sequences are depicted in the table. The red sequence is a tag and used as a template for the second round PCR.

| DMR/ Gene | Chromosomal location | Primer | Sequence 5´-3´ | Length Target [bp] | Annealing Temp. [°C] | Analyzed CpGs |
|---|---|---|---|---|---|---|
| **PTSD cohort** | | | | | | |
| **A) Candidate Genes** | | | | | | |
| *NR3C1* | chr5:142.783.541-142.783.911 | forward | CTTGCTTCCTGGCACGAGGGGGGTAGATTTGGTTTTTT | 371 | 56.9 | 42 |
| | | reverse | CAGGAAACAGCTATGACTCCCTTCCCTAAAACCTC | | | |
| *SLC6A4 Promoter* | chr17:28.562.939-28.563.283 | forward | CTTGCTTCCTGGCACGAGTAGGAGGGGAGGGATTTT | 345 | 59.4 | 23 |
| | | reverse | CAGGAAACAGCTATGACAAACCTCTAAACTAAACTCACATC | | | |
| | chr17:28.562.574-28.562.952 | forward | CTTGCTTCCTGGCACGAGGGGAAGAAGGTTTGGAAAGA | 379 | 59.4 | 42 |
| | | reverse | CAGGAAACAGCTATGACTCCCTCCCCTCCTAACTCTAA | | | |
| | chr17:28.562.328-28.562.682 | forward | CTTGCTTCCTGGCACGAGTTTTTAAGGGTTTTTAAGAGGTTGTAAAGT | 355 | 60.0 | 17 |
| | | reverse | CAGGAAACAGCTATGACAAACCAACCCCCCTACCCAACCC | | | |
| *OXTR Enhancer* | chr3:8.799.262-8.799.615 | forward | CTTGCTTCCTGGCACGAGTTGTGGGTAGGAGTAGGATTTTA | 354 | 59.4 | 5 |
| | | reverse | CAGGAAACAGCTATGACTTCTCATCCTAAATCTAAAAATCACTT | | | |
| | chr3:8.800.371-8.800.739 | forward | CTTGCTTCCTGGCACGAGAGTGTAAGGTTTGGGTGAA | 369 | 54.7 | 11 |
| | | reverse | CAGGAAACAGCTATGACATCTAAAATAAAACCCCAAAAATT | | | |
| *FKBP5* | chr6:35.558.322-35.558.593 272 | forward | CTTGCTTCCTGGCACGAGTTTTGGGTTGAGGATAGAAAGG | 272 | 56 | 5 |
| | | reverse | CAGGAAACAGCTATGACATCCAAAACAACTAACAAATTCTCT | | | |
| **B) New Targets from WGBS** | | | | | | |
| DMR-4 *ADORA1* | chr1:203.097.582-203.097.833 | forward | CTTGCTTCCTGGCACGAGTTGTGTATAGGGGTGGGTAGA | 252 | 61.5 | 7 |
| | | reverse | CAGGAAACAGCTATGACACCCTATAATATATCCCACTTATCAC | | | |
| DMR-21 *TSPAN9* | chr12:3.370.787-3.371.092 | forward | CTTGCTTCCTGGCACGAGAATGGGGATGTTTAGTTAGTGTTATTTAG | 306 | 61.5 | 6 |
| | | reverse | CAGGAAACAGCTATGACACACACAAAAAATAAACCTACTACTTTTC | | | |

| | | | | | | |
|---|---|---|---|---|---|---|
| DMR-10 *RPS6KA2* | chr6:166.996.725-166.997.045 | forward | CTTGCTTCCTGGCACGAGTGGGGAATTTTGGTGTAGATATGA | 321 | 61.5 | 8 |
| | | reverse | CAGGAAACAGCTATGACCACTACCTCATCCAAAACCCTAAT | | | |
| DMR-1 | chr1:149.233.322-149.233.624 | forward | CTTGCTTCCTGGCACGAGTGAAGTTTTTTTGTAGGTTATAGGGAAGGG | 303 | 61.5 | 14 |
| | | reverse | CAGGAAACAGCTATGACCTACCCAATCTTTCTCTTTCTTAAT | | | |
| **AN cohort** | | | | | | |
| **C) New Targets from WGBS** | | | | | | |
| DMR-1 *GLB1L* | chr2:220.107.940-220.108.293 | forward | CTTGCTTCCTGGCACGAGGAAAGTAGTGTAGGTTGTTAGA | 354 | 55 | 22 |
| | | reverse | CAGGAAACAGCTATGACAACTCCCAAAAAACTATCC | | | |
| DMR-3 | chr3:96.495.592-96.495.821 | forward | CTTGCTTCCTGGCACGAGAAAAGGTTTTAATGTAGAATAAAAAT | 230 | 52 | 28 |
| | | reverse | CAGGAAACAGCTATGACTAACAAAAAAATTATACAAACTCC | | | |
| DMR-7 *FAM50B* | chr6:3.849.948-3.850.193 | forward | CTTGCTTCCTGGCACGAGGAGGYGTAGAGTTGAGTATTTTT | 246 | 56 | 19 |
| | | reverse | CAGGAAACAGCTATGACAAAACCTCTTATCCACCTA | | | |
| DMR-11 *MEST* | chr7: 130.132.730-130.133.038 | forward | CTTGCTTCCTGGCACGAGTGTAGGATTTTTAGAATTTTAGT | 309 | 56 | 22 |
| | | reverse | CAGGAAACAGCTATGACCAACATAACAATTTAATCACATC | | | |
| DMR-13 *ERLIN2* | chr8:37.605.428-37.605.657 | forward | CTTGCTTCCTGGCACGAGTAGTAGTATATATGGAGGGGTTTTT | 230 | 50 | 16 |
| | | reverse | CAGGAAACAGCTATGACTAAAAATATTAATAAATACCATTTAAATTA | | | |
| DMR-16 *EXD3* | chr9:140.311.984-140.312.340 | forward | CTTGCTTCCTGGCACGAGGGGTTTTGGTAGTTTTTTTT | 357 | 56 | 44 |
| | | reverse | CAGGAAACAGCTATGACACTCCTCAAATCCTCAAACTCTATC | | | |
| DMR-22 *SNURF/SNRPN* | chr15:25.200.594-25.200.895 | forward | CTTGCTTCCTGGCACGAGGGGATTAGTGTATAGGGATTTTAGG | 302 | 60 | 19 |
| | | reverse | CAGGAAACAGCTATGACCTTCCCCCTACCTCCCAA | | | |
| DMR-28 *HSPA12B* | chr20:3.732.639-3.732.992 | forward | CTTGCTTCCTGGCACGAGGTTTAGTTTYGAGTTTGAGTT | 354 | 55 | 41 |
| | | reverse | CAGGAAACAGCTATGACCAATCTCTAAATATATCCCCACC | | | |

Length of the amplicons is stated without tags.

2.PCR: The second round PCR reaction contained 1 µl of PCR product of the first round. 0.2 µM primers and 5 µl GoTaq® G2 Hot Start Master Mixes (Promega. Fitchburg. WI. USA) in a total volume of 10 µl. The standard amplification protocol included an initial denaturation step for 2 min at 95 °C. followed by 35 cycles of melting at 94 °C for 30 s. annealing and extension at 72 °C for 1 min. Followed by an additional extension at 72 °C for 10 min at the end of the 35 cycles. All PCRs included nontemplate controls. Different combinations of Illumina adapters N701-N712 and [N/S/E]501-[N/S/E]508 and [N/S/E]517) were used. introducing index and variable sequences. sequencing primer binding sites and regions complementary to the flow cell Oligos which are critical for cluster generation. The combination of index and variable sequences in forward and reverse primers are unique to each set of primer pair and serve as identifiers for each study subject. As they are specific to a given sample library they enable multiple sequences to be sequenced together and are used for de-multiplexing during data analysis to assign individual sequence reads to the correct sample during final data analysis.

#### 2.3 S3 Table. Linear regression with pre-treatment mean DNA methylation and severity of baseline PTSD symptoms

| A) Candidate Genes | Variable | B | β | s.e. | p |
|---|---|---|---|---|---|
| ***NR3C1*** | α | 0.010 | | 0.001 | |
| | PCL-pre | $-4.907 \times 10^{-6}$ | -0.032 | 0.000 | 0.820 |
| | $R^2$ | 0.001 | | | |
| | corr. $R^2$ | -0.018 | | | |
| | F(df=1;52) | 0.053 | | | |
| ***SLC6A4*** | α | 0.061 | | 0.005 | |
| | PCL-pre | $7.945 \times 10^{-5}$ | 0.119 | 0.000 | 0.376 |
| | $R^2$ | 0.014 | | | |
| | corr. $R^2$ | -0.004 | | | |
| | F(df=1;56) | 0.798 | | | |
| ***OXTR*** | α | 0.908 | | 0.010 | |
| | PCL-pre | 0.000 | 0.187 | 0.000 | 0.160 |
| | $R^2$ | 0.035 | | | |
| | corr. $R^2$ | 0.018 | | | |
| | F(df=1;56) | 2.023 | | | |
| ***FKBP5*** | α | 0.875 | | 0.010 | |
| | PCL-pre | 0.000 | 0.208 | 0.000 | 0.118 |
| | $R^2$ | 0.043 | | | |
| | corr. $R^2$ | 0.026 | | | |
| | F(df=1;56) | 2.523 | | | |
| **B) New Targets from WGBS** | Variable | **B** | ***β*** | **s.e.** | **p** |
| ***ADORA1*** | α | 0.442 | | 0.040 | |
| | PCL-pre | -0.001 | -0.133 | 0.001 | 0.319 |
| | $R^2$ | 0.018 | | | |
| | corr. $R^2$ | 0.000 | | | |
| | F(df=1;56) | 1.010 | | | |
| ***TSPAN9*** | α | 0.508 | | 0.083 | |
| | PCL-pre | 0.002 | 0.145 | 0.001 | 0.279 |
| | $R^2$ | 0.021 | | | |
| | corr. $R^2$ | 0.003 | | | |
| | F(df=1;56) | 1.196 | | | |
| ***RSP6KA2*** | α | 0.499 | | 0.046 | |
| | PCL-pre | 0.000 | -0.018 | 0.001 | 0.895 |
| | $R^2$ | 0.000 | | | |
| | corr. $R^2$ | -0.018 | | | |
| | F(df=1;55) | 0.018 | | | |
| **DMR-1** | α | 0.714 | | 0.044 | |
| | PCL-pre | 0.000 | 0.043 | 0.001 | 0.746 |
| | $R^2$ | 0.002 | | | |
| | corr. $R^2$ | -0.016 | | | |
| | F(df=1;56) | 0.106 | | | |

#### 2.4 S4 Table. Mean DNA methylation levels at single CpG sites

##### A) Candidate Genes

**NR3C1** Location: chr5:142783911-142783541

| CpG No. | Pre Intervention | Post Intervention | Difference | SD | $F_{(df=1;55)}$ | *p* |
|---|---|---|---|---|---|---|
| CpG_1 | 0.66% | 0.54% | -0.13% | 0.95% | .960 | .331 |
| CpG_2 | 1.05% | 0.89% | -0.16% | 1.01% | 1.432 | .237 |
| CpG_3 | 1.02% | 1.05% | 0.04% | 1.26% | .045 | .833 |
| CpG_4 | 0.70% | 1.00% | 0.30% | 1.19% | 3.646 | .061 |
| CpG_5 | 0.64% | 0.82% | 0.18% | 1.39% | .925 | .340 |
| CpG_6 | 0.96% | 0.70% | -0.27% | 1.31% | 2.327 | .133 |
| CpG_7 | 0.80% | 1.02% | 0.21% | 1.38% | 1.341 | .252 |
| CpG_8 | 0.48% | 0.50% | 0.02% | 0.92% | .021 | .886 |
| CpG_9 | 0.59% | 0.55% | -0.04% | 0.93% | .082 | .776 |
| CpG_10 | 0.88% | 0.73% | -0.14% | 1.14% | .887 | .350 |
| CpG_11 | 0.71% | 0.55% | -0.16% | 1.02% | 1.382 | .245 |
| CpG_12 | 0.68% | 0.93% | 0.25% | 1.30% | 2.081 | .155 |
| CpG_13 | 1.13% | 1.36% | 0.23% | 1.61% | 1.169 | .284 |
| CpG_14 | 0.66% | 0.61% | -0.05% | 1.13% | .125 | .725 |
| CpG_15 | 0.79% | 0.61% | -0.18% | 1.28% | 1.089 | .301 |
| CpG_16 | 0.68% | 0.50% | -0.18% | 0.94% | 2.037 | .159 |
| CpG_17 | 0.95% | 1.18% | 0.23% | 1.29% | 1.805 | .185 |
| CpG_18 | 0.98% | 1.13% | 0.14% | 1.27% | .707 | .404 |
| CpG_19 | 0.86% | 0.93% | 0.07% | 1.20% | .197 | .659 |
| CpG_20 | 0.75% | 0.73% | -0.02% | 0.92% | .021 | .886 |
| CpG_21 | 0.66% | 0.57% | -0.09% | 1.08% | .380 | .540 |
| CpG_22 | 1.09% | 1.07% | -0.02% | 1.38% | .009 | .923 |
| CpG_23 | 0.96% | 0.73% | -0.23% | 1.06% | 2.678 | .107 |
| CpG_24 | 1.21% | 1.41% | 0.20% | 1.18% | 1.547 | .219 |
| CpG_25 | 1.63% | 1.11% | -0.52% | 1.43% | 7.376 | .009 |
| CpG_26 | 1.11% | 1.20% | 0.09% | 1.32% | .254 | .616 |
| CpG_27 | 1.20% | 1.34% | 0.14% | 1.46% | .538 | .466 |
| CpG_28 | 1.05% | 0.98% | -0.07% | 1.22% | .192 | .663 |
| CpG_29 | 1.02% | 0.91% | -0.11% | 1.07% | .558 | .458 |
| CpG_30 | 0.88% | 0.80% | -0.07% | 1.06% | .255 | .616 |
| CpG_31 | 1.36% | 1.02% | -0.34% | 1.63% | 2.419 | .126 |
| CpG_32 | 0.80% | 0.59% | -0.21% | 1.19% | 1.827 | .182 |
| CpG_33 | 0.66% | 0.70% | 0.04% | 0.99% | .073 | .788 |
| CpG_34 | 1.30% | 1.32% | 0.02% | 1.48% | .008 | .929 |
| CpG_35 | 0.59% | 0.66% | 0.07% | 1.02% | .272 | .604 |
| CpG_36 | 0.64% | 0.73% | 0.09% | 1.25% | .284 | .596 |
| CpG_37 | 1.46% | 1.34% | -0.13% | 1.27% | .546 | .463 |
| CpG_38 | 1.13% | 1.04% | -0.09% | 1.35% | .244 | .623 |
| CpG_39 | 2.05% | 1.96% | -0.09% | 1.82% | .135 | .715 |
| CpG_40 | 1.61% | 1.36% | -0.25% | 1.28% | 2.127 | .150 |
| CpG_41 | 1.04% | 1.14% | 0.11% | 1.29% | .387 | .536 |
| CpG_42 | 0.91% | 1.21% | 0.30% | 1.19% | 3.646 | .061 |

**SLC6A4** Location: chr17:28563283-28562328

| CpG No. | Pre Intervention | Post Intervention | Difference | SD | $F_{(df=1;59)}$ | *p* |
|---|---|---|---|---|---|---|
| CpG_03 | 5.23% | 5.32% | 0.08% | 2.59% | .062 | .804 |

| | | | | | | |
|---|---|---|---|---|---|---|
| CpG_02 | 1.98% | 2.27% | 0.28% | 1.38% | 2.533 | .117 |
| CpG_01 | 2.38% | 2.48% | 0.10% | 1.35% | .330 | .568 |
| CpG_1 | 4.62% | 4.72% | 0.10% | 2.75% | .079 | .779 |
| CpG_2 | 1.62% | 1.80% | 0.18% | 1.24% | 1.308 | .257 |
| CpG_3 | 1.73% | 1.58% | -0.15% | 1.33% | .768 | .384 |
| CpG_4 | 3.00% | 2.70% | -0.30% | 1.77% | 1.726 | .194 |
| CpG_5 | 1.00% | 0.80% | -0.20% | 0.94% | 2.744 | .103 |
| CpG_6 | 0.85% | 0.85% | 0.00% | 0.97% | 0.000 | 1.000 |
| CpG_7 | 1.48% | 1.30% | -0.18% | 1.32% | 1.155 | .287 |
| CpG_8 | 1.57% | 1.63% | 0.07% | 1.34% | .149 | .701 |
| CpG_9 | 1.73% | 1.72% | -0.02% | 1.14% | .013 | .910 |
| CpG_10 | 2.45% | 2.50% | 0.05% | 1.63% | .056 | .813 |
| CpG_11 | 2.12% | 2.35% | 0.23% | 1.78% | 1.032 | .314 |
| CpG_12 | 2.85% | 2.68% | -0.17% | 1.62% | .637 | .428 |
| CpG_13 | 1.43% | 1.52% | 0.08% | 1.18% | .298 | .587 |
| CpG_14 | 2.03% | 2.15% | 0.12% | 1.62% | .313 | .578 |
| CpG_15 | 2.42% | 2.23% | -0.18% | 1.72% | .680 | .413 |
| CpG_16 | 1.72% | 2.02% | 0.30% | 1.75% | 1.764 | .189 |
| CpG_17 | 3.53% | 4.08% | 0.55% | 2.00% | 4.521 | .038 |
| CpG_18 | 1.37% | 1.58% | 0.22% | 1.62% | 1.078 | .303 |
| CpG_19 | 1.58% | 1.82% | 0.23% | 2.68% | .454 | .503 |
| CpG_20 | 1.07% | 1.33% | 0.27% | 2.28% | .818 | .369 |
| CpG_21 | 1.02% | 1.05% | 0.03% | 1.19% | .047 | .829 |
| CpG_22 | 1.85% | 1.83% | -0.02% | 1.66% | .006 | .938 |
| CpG_23 | 2.15% | 2.12% | -0.03% | 1.33% | .038 | .846 |
| CpG_24 | 1.90% | 1.68% | -0.22% | 1.60% | 1.107 | .297 |
| CpG_25 | 2.92% | 2.70% | -0.22% | 1.73% | .943 | .335 |
| CpG_26 | 2.02% | 1.87% | -0.15% | 1.61% | .518 | .474 |
| CpG_27 | 2.92% | 2.67% | -0.25% | 2.04% | .141 | .709 |
| CpG_28 | 1.98% | 1.82% | -0.17% | 1.42% | .831 | .366 |
| CpG_29 | 5.63% | 5.95% | 0.32% | 2.43% | 1.023 | .316 |
| CpG_30 | 2.62% | 2.02% | -0.60% | 1.71% | 7.392 | .009 |
| CpG_31 | 2.08% | 1.77% | -0.32% | 1.51% | 2.630 | .110 |
| CpG_32 | 3.53% | 3.00% | -0.53% | 2.37% | 3.043 | .086 |
| CpG_33 | 1.55% | 1.70% | 0.15% | 1.67% | .487 | .488 |
| CpG_34 | 1.62% | 1.85% | 0.23% | 1.38% | 1.710 | .196 |
| CpG_35 | 2.08% | 1.98% | -0.10% | 1.72% | .202 | .655 |
| CpG_36 | 3.23% | 2.70% | -0.53% | 1.88% | 4.819 | .032 |
| CpG_37 | 2.27% | 2.32% | 0.05% | 1.60% | .059 | .809 |
| CpG_38 | 1.67% | 1.75% | 0.08% | 1.32% | .240 | .626 |
| CpG_39 | 2.80% | 2.45% | -0.35% | 1.69% | 2.587 | .113 |
| CpG_40 | 3.15% | 2.83% | -0.32% | 2.13% | 1.330 | .254 |
| CpG_41 | 1.25% | 1.07% | -0.18% | 1.10% | 1.676 | .200 |
| CpG_42 | 1.38% | 1.38% | 0.00% | 1.18% | 0.000 | 1.000 |
| CpG_43 | 1.73% | 1.83% | 0.10% | 1.54% | .254 | .616 |
| CpG_44 | 1.90% | 1.73% | -0.17% | 1.39% | .860 | .358 |
| CpG_45 | 2.45% | 2.42% | -0.03% | 1.51% | .029 | .865 |
| CpG_46 | 3.77% | 3.88% | 0.12% | 1.97% | .211 | .648 |
| CpG_47 | 3.23% | 3.08% | -0.15% | 1.91% | .369 | .546 |
| CpG_48 | 6.42% | 6.10% | -0.32% | 2.75% | .794 | .376 |
| CpG_49 | 7.67% | 7.40% | -0.27% | 3.19% | .418 | .520 |
| CpG_50 | 10.15% | 10.48% | 0.33% | 3.29% | .617 | .435 |
| CpG_51 | 9.92% | 9.52% | -0.40% | 3.93% | .622 | .433 |
| CpG_52 | 5.25% | 5.05% | -0.20% | 2.20% | .496 | .484 |
| CpG_53 | 10.97% | 10.32% | -0.65% | 3.37% | 2.227 | .141 |
| CpG_54 | 13.27% | 13.43% | 0.17% | 3.52% | .135 | .715 |
| CpG_55 | 19.32% | 18.52% | -0.80% | 4.67% | 1.760 | .190 |
| CpG_56 | 5.23% | 5.37% | 0.13% | 2.76% | .140 | .709 |
| CpG_57 | 10.62% | 10.95% | 0.33% | 4.10% | .397 | .531 |
| CpG_58 | 12.45% | 11.92% | -0.53% | 4.38% | .889 | .350 |

| | | | | | | |
|---|---|---|---|---|---|---|
| CpG_59 | 13.63% | 13.85% | 0.22% | 3.33% | .253 | .617 |
| CpG_60 | 14.37% | 14.57% | 0.20% | 4.28% | .131 | .719 |
| CpG_61 | 3.58% | 3.60% | 0.02% | 2.13% | .004 | .952 |
| CpG_62 | 1.08% | 1.20% | 0.12% | 1.11% | .668 | .417 |
| CpG_63 | 3.65% | 4.05% | 0.40% | 2.37% | 1.704 | .197 |
| CpG_64 | 3.67% | 3.55% | -0.12% | 2.20% | .168 | .683 |
| CpG_65 | 3.93% | 4.48% | 0.55% | 2.59% | 2.698 | .106 |
| CpG_66 | 18.27% | 19.23% | 0.97% | 3.88% | 3.717 | .059 |
| CpG_67 | 24.67% | 24.23% | -0.43% | 5.26% | .407 | .526 |
| CpG_69 | 29.72% | 31.30% | 1.58% | 5.55% | 4.885 | .031 |
| CpG_70 | 12.17% | 13.20% | 1.03% | 2.80% | 8.183 | .006 |
| CpG_71 | 13.80% | 14.07% | 0.27% | 3.75% | .304 | .583 |
| CpG_72 | 10.23% | 9.63% | -0.60% | 3.51% | 1.750 | .191 |
| CpG_73 | 10.67% | 10.77% | 0.10% | 3.74% | .043 | .836 |
| CpG_74 | 19.82% | 20.20% | 0.38% | 5.19% | .327 | .570 |
| CpG_75 | 21.38% | 21.50% | 0.12% | 5.17% | .031 | .862 |
| CpG_77 | 11.90% | 12.68% | 0.78% | 3.72% | 2.661 | .108 |
| CpG_78 | 8.12% | 8.70% | 0.58% | 2.92% | 2.397 | .127 |
| CpG_79 | 12.87% | 14.03% | 1.17% | 3.84% | 5.549 | .022 |
| CpG_80 | 29.75% | 30.80% | 1.05% | 4.97% | 2.679 | .107 |
| CpG_81 | 41.18% | 41.90% | 0.72% | 5.62% | .975 | .327 |

***OXTR* Enhancer** Location: chr3:8798082-8800976

| CpG No. | Pre Intervention | Post Intervention | Difference | SD | $F_{(df=1;59)}$ | *p* |
|---|---|---|---|---|---|---|
| CpG_5 | 93.15% | 93.38% | 0.23% | 4.14% | .190 | .664 |
| CpG_6 | 95.72% | 95.82% | 0.10% | 7.10% | .012 | .913 |
| CpG_7 | 86.30% | 84.83% | -1.47% | 6.66% | 2.912 | .093 |
| CpG_8 | 95.37% | 95.82% | 0.45% | 3.18% | 1.205 | .277 |
| CpG_9 | 88.67% | 88.45% | -0.22% | 5.73% | .086 | .771 |
| CpG_24 | 93.00% | 93.68% | 0.68% | 4.79% | 1.222 | .274 |
| CpG_25 | 82.98% | 81.70% | -1.28% | 5.97% | 2.776 | .101 |
| CpG_26 | 94.02% | 93.07% | -0.95% | 3.65% | 4.071 | .048 |
| CpG_28 | 96.50% | 96.40% | -0.10% | 2.67% | .084 | .772 |
| CpG_29 | 93.08% | 93.53% | 0.45% | 3.06% | 1.297 | .259 |
| CpG_30 | 96.62% | 96.42% | -0.20% | 2.18% | .506 | .479 |
| CpG_31 | 92.45% | 93.30% | 0.85% | 3.88% | 2.875 | .095 |
| CpG_32 | 95.37% | 94.78% | -0.58% | 3.57% | 1.601 | .211 |
| CpG_33 | 89.87% | 89.05% | -0.82% | 5.32% | 1.416 | .239 |
| CpG_34 | 90.42% | 90.12% | -0.30% | 3.78% | .378 | .541 |
| CpG_35 | 90.80% | 89.75% | -1.05% | 6.45% | 1.591 | .212 |

**FKBP5** Location: chr6:35558322-35558593

| CpG No. | Pre Intervention | Post Intervention | Difference | SD | $F_{(df=1;59)}$ | *p* |
|---|---|---|---|---|---|---|
| CpG_1 | 78.93% | 79.12% | 0.18% | 2.59% | .300 | .586 |
| CpG_2 | 97.15% | 97.23% | 0.08% | 1.00% | .420 | .520 |
| CpG_3 | 97.83% | 97.95% | 0.12% | 1.15% | .616 | .436 |
| CpG_4 | 80.98% | 81.05% | 0.07% | 2.65% | .038 | .846 |
| CpG_5 | 89.92% | 90.07% | 0.15% | 2.72% | .183 | .671 |

#### B) New Targets from WGBS

***ADORA1*** Location: chr1:203097582-203097833

| CpG No. | Pre Intervention | Post Intervention | Difference | SD | $F_{(df=1;59)}$ | *p* |
|---|---|---|---|---|---|---|
| CpG_1 | 19.57% | 18.80% | -0.77% | 4.64% | 1.637 | .206 |
| CpG_2 | 53.35% | 53.52% | 0.17% | 5.25% | .061 | .807 |
| CpG_3 | 40.30% | 42.53% | 2.23% | 9.15% | 3.574 | .064 |
| CpG_4 | 41.62% | 42.67% | 1.05% | 8.27% | .967 | .329 |
| CpG_5 | 48.85% | 48.55% | -0.30% | 7.95% | .085 | .771 |
| CpG_6 | 26.32% | 27.92% | 1.60% | 6.74% | 3.378 | .071 |
| CpG_7 | 52.08% | 53.97% | 1.88% | 8.87% | 2.704 | .105 |

***TSPAN9*** Location: chr12:3370787-3371092

| CpG No. | Pre Intervention | Post Intervention | Difference | SD | $F_{(df=1;59)}$ | *p* |
|---|---|---|---|---|---|---|
| CpG_1 | 60.28% | 55.57% | -4.72% | 18.91% | 3.732 | .058 |
| CpG_2 | 83.42% | 82.65% | -0.77% | 19.53% | .092 | .762 |
| CpG_3 | 52.83% | 51.33% | -1.50% | 17.32% | .450 | .505 |
| CpG_4 | 43.80% | 40.70% | -3.10% | 17.02% | 1.991 | .163 |
| CpG_5 | 63.57% | 59.93% | -3.63% | 16.11% | 3.053 | .086 |
| CpG_6 | 54.73% | 51.73% | -3.00% | 15.50% | 2.249 | .139 |

***RPS6KA2*** Location: chr6:166997045-166996725

| CpG No. | Pre Intervention | Post Intervention | Difference | SD | $F_{(df=1;58)}$ | *p* |
|---|---|---|---|---|---|---|
| CpG_1 | 79.80% | 81.19% | 1.39% | 6.56% | 2.646 | .109 |
| CpG_2 | 56.85% | 57.17% | 0.32% | 7.14% | .120 | .730 |
| CpG_3 | 67.66% | 68.07% | 0.41% | 6.66% | .220 | .641 |
| CpG_4 | 36.81% | 36.31% | -0.51% | 7.23% | .292 | .591 |
| CpG_5 | 23.90% | 23.53% | -0.37% | 6.90% | .173 | .679 |
| CpG_6 | 52.39% | 52.56% | 0.17% | 6.05% | .046 | .831 |
| CpG_7 | 38.56% | 37.71% | -0.85% | 7.11% | .837 | .364 |
| CpG_8 | 38.15% | 37.31% | -0.85% | 7.63% | .728 | .397 |

**DMR-1** Location: chr1:149233322-149233624

| CpG No. | Pre Intervention | Post Intervention | Difference | SD | $F_{(df=1;59)}$ | *p* |
|---|---|---|---|---|---|---|
| CpG_2 | 54.63% | 55.15% | 0.52% | 6.19% | .418 | .521 |
| CpG_3 | 43.47% | 43.05% | -0.42% | 5.57% | .336 | .564 |
| CpG_4 | 64.43% | 63.37% | -1.07% | 6.22% | 1.765 | .189 |
| CpG_6 | 68.55% | 68.70% | 0.15% | 6.14% | .036 | .850 |
| CpG_7 | 64.05% | 64.02% | -0.03% | 6.63% | .002 | .969 |
| CpG_8 | 75.40% | 74.70% | -0.70% | 5.81% | .871 | .355 |
| CpG_9 | 82.80% | 82.72% | -0.08% | 4.80% | .018 | .893 |
| CpG_10 | 57.87% | 57.67% | -0.20% | 7.18% | .047 | .830 |
| CpG_11 | 78.22% | 77.47% | -0.75% | 5.54% | 1.102 | .298 |
| CpG_12 | 77.18% | 77.53% | 0.35% | 5.93% | .209 | .649 |
| CpG_13 | 81.43% | 81.75% | 0.32% | 5.13% | .229 | .634 |
| CpG_14 | 87.35% | 87.50% | 0.15% | 4.99% | .054 | .817 |
| CpG_15 | 90.60% | 90.33% | -0.27% | 3.80% | .296 | .589 |
| CpG_16 | 92.42% | 92.73% | 0.32% | 3.72% | .436 | .512 |

#### 2.5 S5 Table. WGBS descriptive information

| Cohort | Sample (ENA) | before/after | Conversion rate | Mapping efficiency | Duplication Rate | Coverage | Mean methylation |
|---|---|---|---|---|---|---|---|
| PTSD | 47.1 (K002000217_85588) | before | 1 | 1 | 0.21 | 9.11 | 0.72 |
| PTSD | 47.2 (K002000217_85589) | after | 1 | 1 | 0.21 | 8.33 | 0.72 |
| PTSD | 43.1(K002000217_85590) | before | 1 | 1 | 0.22 | 8.19 | 0.72 |
| PTSD | 43.2 (K002000217_85591) | after | 1 | 1 | 0.22 | 8.07 | 0.72 |
| Anorexia | AN1-pre (AS-222872-LR-34031) | before | 1 | 1 | 0.19 | 21.15 | 0.76 |
| Anorexia | AN1-post (AS-222873-LR-34032) | after | 1 | 1 | 0.18 | 22.54 | 0.76 |
| Anorexia | AN2-pre (AS-222875-LR-34033) | before | 1 | 1 | 0.17 | 21.13 | 0.76 |
| Anorexia | AN2-post (AS-222876-LR-34034) | after | 1 | 1 | 0.15 | 21.17 | 0.76 |
| Anorexia | AN3-pre (AS-222877-LR-34035) | before | 1 | 1 | 0.20 | 18.75 | 0.75 |
| Anorexia | AN3-post (AS-241990-LR-35191) | after | 1 | 1 | 0.17 | 18.00 | 0.74 |

#### 2.6 S6 Table. DMRs in the PTSD cohort identified with camel

| # | chrom | start | stop | cpg | mean_meth_diff | gene | cgi | overlap metilene | DBS |
|---|---|---|---|---|---|---|---|---|---|
| 1 | 1 | 149233364 | 149233394 | 4 | 0.31 | | | | yes |
| 2 | 1 | 229119599 | 229120672 | 8 | 0.30 | | | | |
| 3 | 1 | 3685641 | 3685665 | 4 | -0.30 | *CCDC27* | | | |
| 4 | 1 | 203097628 | 203097776 | 7 | -0.32 | *ADORA1* | | | yes |
| 5 | 2 | 178034563 | 178034833 | 5 | 0.30 | | | | |
| 6 | 2 | 88302827 | 88302847 | 4 | -0.40 | | | | |
| 7 | 2 | 104445362 | 104447294 | 8 | -0.37 | | | | |
| 8 | 4 | 132984311 | 132984336 | 4 | -0.30 | | | | |
| 9 | 5 | 4512352 | 4512371 | 4 | 0.32 | | | | |
| 10 | 6 | 166996807 | 166996856 | 4 | 0.31 | *RPS6KA2* | | | yes |
| 11 | 6 | 28863695 | 28863746 | 4 | -0.41 | | | yes | |
| 12 | 7 | 1595247 | 1595280 | 4 | 0.34 | *TMEM184A* | | | |
| 13 | 7 | 101514184 | 101514315 | 5 | 0.31 | *CUX1* | | | |
| 14 | 7 | 148768873 | 148768932 | 7 | -0.39 | *ZNF786* | yes | | |
| 15 | 8 | 35549566 | 35549812 | 4 | 0.31 | *UNC5D* | | | |
| 16 | 8 | 141109357 | 141109414 | 5 | 0.32 | *TRAPPC9* | yes | | |
| 17 | 10 | 131052326 | 131052347 | 4 | -0.31 | | | | |
| 18 | 11 | 1892538 | 1892592 | 4 | 0.38 | *LSP1* | yes | | |
| 19 | 11 | 45951791 | 45951817 | 4 | -0.33 | *PHF21A* | | | |
| 20 | 12 | 130615735 | 130615819 | 4 | 0.31 | | | | |
| 21 | 12 | 3370834 | 3371031 | 6 | -0.36 | *TSPAN9* | | | yes |
| 22 | 12 | 133181087 | 133181120 | 4 | -0.40 | *LRCOL1* | | | |
| 23 | 13 | 41496184 | 41496213 | 5 | -0.33 | | yes | | |
| 24 | 15 | 88874573 | 88874627 | 4 | -0.33 | | | | |
| 25 | 16 | 75031951 | 75032022 | 7 | 0.30 | | | | |
| 26 | 16 | 81924284 | 81924435 | 5 | 0.38 | *PLCG2* | | | |
| 27 | 20 | 57426931 | 57427018 | 11 | 0.30 | *GNAS* | | | |
| 28 | 20 | 57430980 | 57431013 | 4 | -0.38 | *GNAS* | yes | | |
| 29 | 22 | 19086801 | 19086831 | 4 | 0.35 | *DGCR2* | | | |
| 30 | X | 74144931 | 74144954 | 4 | 0.37 | *KIAA2022* | yes | | |
| 31 | X | 102879244 | 102879668 | 6 | 0.37 | | | | |
| 32 | X | 119149621 | 119149673 | 8 | 0.35 | | yes | | |
| 33 | X | 34836096 | 34836268 | 4 | -0.30 | | | | |

#### 2.7 S7 Table. DMRs in the PTSD cohort identified with metilene

| # | #chr | start | stop | q-value | mean methylation difference | #CpGs | p (MWU) | p (2D KS) | mean g1 | mean g2 | cgi | gene | overlap camel |
|---|---|---|---|---|---|---|---|---|---|---|---|---|---|
| 1 | 6 | 28863663 | 28863803 | 0.0270 | 36.36 | 11 | 1.30E-08 | 3.90E-09 | 55.682 | 19.318 | | | yes |
| 2 | 6 | 32847513 | 32847758 | 0.0450 | 33.72 | 16 | 1.40E-10 | 6.70E-09 | 86.875 | 53.156 | yes | | |
| 3 | 8 | 105379654 | 105379756 | 0.0140 | 39.54 | 12 | 3.70E-09 | 2.10E-09 | 72.417 | 32.875 | yes | | |
| 4 | 9 | 140312096 | 140312262 | 0.0037 | 31.76 | 25 | 1.30E-13 | 5.40E-10 | 83.760 | 52.000 | yes | *EXD3* | |

#### 2.8 S8 Table. DMRs in the AN cohort identified with camel

| # | chrom | start | stop | cpg | mean_meth_diff | gene | cgi | overlap metilene | DBS |
|---|---|---|---|---|---|---|---|---|---|
| 1 | 2 | 220108022 | 220108240 | 15 | -0.20 | *GLB1L* | yes | | yes |
| 2 | 3 | 122631656 | 122631863 | 19 | 0.20 | *SEMA5B* | yes | | |
| 3 | 3 | 96495654 | 96495787 | 25 | -0.24 | | yes | yes | yes |
| 4 | 3 | 128372270 | 128372392 | 12 | -0.22 | *RPN1* | | | |
| 5 | 5 | 23951406 | 23951556 | 23 | 0.21 | | | | |
| 6 | 5 | 131607580 | 131607728 | 15 | 0.23 | *PDLIM4* | yes | | |
| 7 | 6 | 3850030 | 3850198 | 16 | 0.25 | *FAM50B* | yes | | yes |
| 8 | 6 | 103780330 | 103780520 | 12 | 0.24 | | | | |
| 9 | 6 | 144329661 | 144329803 | 14 | 0.21 | *PLAGL1* | yes | | |
| 10 | 7 | 50850481 | 50850557 | 10 | 0.21 | *GRB10* | yes | | |
| 11 | 7 | 130132831 | 130132968 | 14 | 0.21 | *MEST* | yes | yes | yes |
| 12 | 7 | 138349196 | 138349306 | 12 | 0.21 | *SVOPL* | yes | yes | |
| 13 | 8 | 37605479 | 37605612 | 13 | 0.27 | *ERLIN2* | | yes | yes |
| 14 | 8 | 145577563 | 145577699 | 12 | -0.21 | *TMEM249* | yes | | |
| 15 | 9 | 121571655 | 121571834 | 22 | 0.28 | | | yes | |
| 16 | 9 | 140312005 | 140312139 | 22 | 0.28 | *EXD3* | yes | yes | yes |
| 17 | 9 | 140311650 | 140311755 | 14 | -0.26 | *EXD3* | yes | | |
| 18 | 10 | 2543763 | 2543863 | 10 | 0.24 | | yes | | |
| 19 | 10 | 134312364 | 134312602 | 11 | 0.23 | | | | |
| 20 | 12 | 297564 | 297660 | 13 | 0.20 | | | | |
| 21 | 14 | 104394643 | 104394730 | 18 | 0.25 | | | | |
| 22 | 15 | 25200653 | 25200867 | 17 | 0.21 | *SNURF/SNRPN* | yes | | yes |
| 23 | 16 | 60558 | 60623 | 18 | -0.24 | | | yes | |
| 24 | 19 | 1423673 | 1423847 | 12 | 0.22 | *DAZAP1* | yes | | |
| 25 | 19 | 50435883 | 50436183 | 13 | 0.22 | *ATF5* | | | |
| 26 | 19 | 54927810 | 54928046 | 22 | 0.21 | *TTYH1* | yes | | |
| 27 | 20 | 3732270 | 3732372 | 17 | 0.23 | *HSPA12B* | yes | | |
| 28 | 20 | 3732712 | 3732826 | 19 | 0.27 | *HSPA12B* | yes | | yes |
| 29 | 20 | 57427046 | 57427278 | 12 | 0.21 | *GNAS* | yes | | |
| 30 | 20 | 57416457 | 57416590 | 14 | -0.23 | *GNAS/GNAS-AS1* | yes | | |
| 31 | X | 47509923 | 47510197 | 27 | 0.21 | *ELK1* | yes | | |
| 32 | X | 128657066 | 128657256 | 16 | 0.28 | *SMARCA1* | yes | | |
| 33 | X | 153599077 | 153599242 | 22 | 0.24 | *FLNA* | yes | | |
| 34 | X | 103411321 | 103411411 | 12 | -0.21 | *FAM199X* | yes | | |
| 35 | X | 119444838 | 119444969 | 13 | -0.21 | *TMEM255A* | yes | | |

#### 2.9 S9 Table. DMRs in the AN cohort identified with metilene

| # | chr | start | stop | q-value | mean methylation difference | #CpGs | p (MWU) | p (2D KS) | mean g1 | mean g2 | cgi | gene | overlap camel | DBS |
|---|---|---|---|---|---|---|---|---|---|---|---|---|---|---|
| 1 | 9 | 140312005 | 140312139 | 3.7E-07 | -27.61 | 22 | 4.2E-14 | 4.3E-14 | 39.712 | 67.318 | yes | *EXD3* | yes | yes |
| 2 | 8 | 37605479 | 37605612 | 0.000039 | -28.76 | 11 | 5.5E-12 | 4.5E-12 | 12.788 | 41.545 | | *ERLIN2* | yes | yes |
| 3 | 16 | 60504 | 60682 | 0.000093 | 21.84 | 29 | 4.2E-14 | 1.1E-11 | 55.126 | 33.287 | | | yes | |
| 4 | 9 | 121571655 | 121571834 | 0.001 | -28.48 | 21 | 4.3E-14 | 1.2E-10 | 43.921 | 72.397 | | | yes | |
| 5 | 7 | 138349215 | 138349321 | 0.003 | -21.06 | 11 | 1.6E-11 | 3.4E-10 | 16.727 | 37.788 | yes | *SVOPL* | yes | |
| 6 | 7 | 130132857 | 130132968 | 0.019 | -21.81 | 12 | 3.3E-11 | 2.2E-09 | 35.250 | 57.056 | yes | *MEST* | yes | yes |
| 7 | 3 | 96495630 | 96495787 | 0.026 | 23.22 | 26 | 8.7E-14 | 2.9E-09 | 73.321 | 50.103 | yes | | yes | yes |
| 8 | X | 153657063 | 153657259 | 0.00038 | 21.71 | 28 | 4.1E-14 | 4.4E-11 | 35.595 | 13.881 | yes | *ATP6AP1* | | |
| 9 | 7 | 105596474 | 105596577 | 0.0021 | -22.90 | 14 | 4.7E-13 | 2.4E-10 | 60.048 | 82.952 | | *CDHR3* | | |
| 10 | X | 46618447 | 46618564 | 0.0043 | 24.73 | 17 | 1E-13 | 4.9E-10 | 33.020 | 8.294 | yes | *SLC9A7* | | |

### 3. Supporting Figures

#### 3.1 S1 Fig. Chromosomal location of analyzed regions of the PTSD cohort

Chromosomal positions of analyzed genomic regions are displayed using graphical output from the UCSC genome browser. All genes are shown in 5´→3´ orientation from left to right. The analyzed DNA regions are shown as red bars. CpG islands with the number of CpGs are shown in green bars.

**A) <u>Candidate Genes</u>**

### *3.1.1 NR3C1*

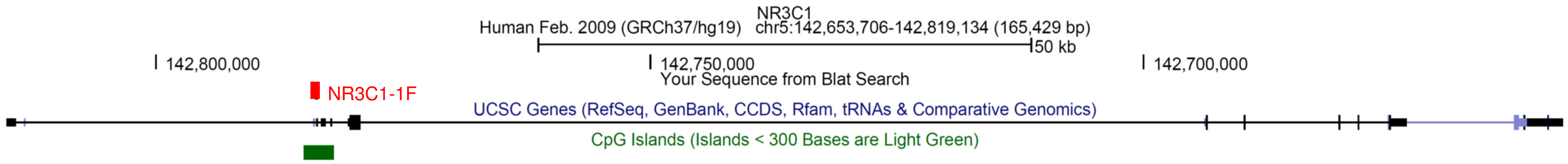

***NR3C1:*** DNA methylation of 42 CpGs in the promoter region of exon 1F of *NR3C1* (Glucocorticoid Receptor) was analyzed (chr5:142.783.541-142.783.911).

##### *3.1.2 SLC6A4*

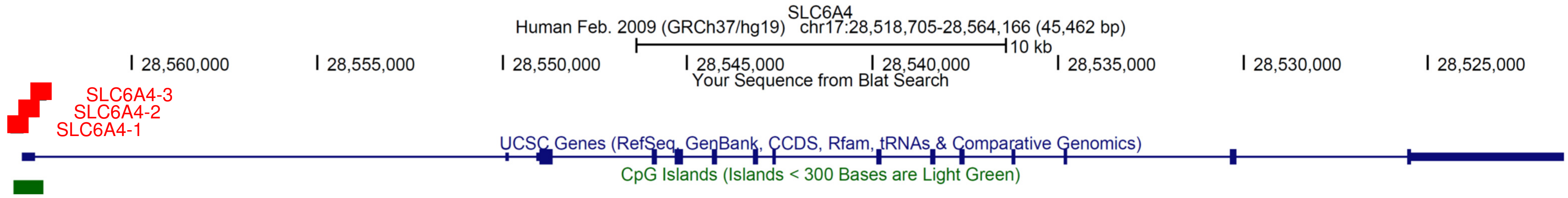

***SLC6A4:*** DNA methylation of the entire CpG island located in the promoter region of the *SLC6A4* (Serotonin Transporter) was analyzed. The CpG island was divided into 3 fragments for sequencing (SLC6A4-1:chr17:28.562.939-28.563.283; SLC6A4-2: chr17:28.562.574-28.562.952; SLC6A4-3: chr17:28.562.328-28.562.682). CpG sites which are affected by SNPs were not included in the statistical analysis (rs25533; rs35206195; and rs56384968).

##### 3.1.3 OXTR

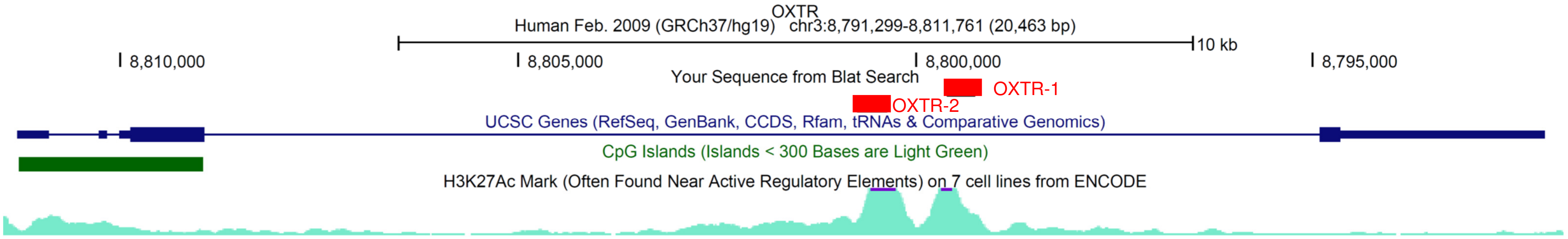

***OXTR:*** 16 CpGs in a putative *OXTR* (Oxytocin Receptor) enhancer region (intron 3) were analyzed (OXTR-1: chr3:8.799.262-8.799.615; OXTR-2: chr3:8.800.371-8.800.739). This region is characterized by histone acetylation (H3K27Ac: turquoise peaks). CpG sites which are affected by SNPs were not analyzed in the statistical analysis (rs2268491 and rs7636061).

##### 3.1.4 FKBP5

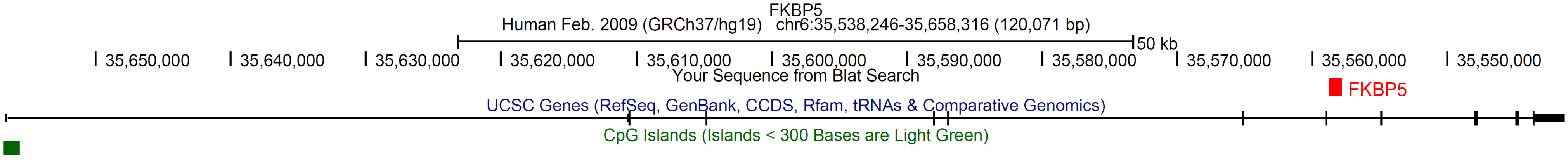

***FKBP5***: DNA methylation analysis was performed on 5 CpGs in intron 7 of *FKBP5* (FKBP Prolyl Isomerase 5) containing a glucocorticoid response element (GRE) (chr6:35.558.322-35.558.593 272).

**B) New Targets from WGBS**

##### 3.1.5 *ADORA1*

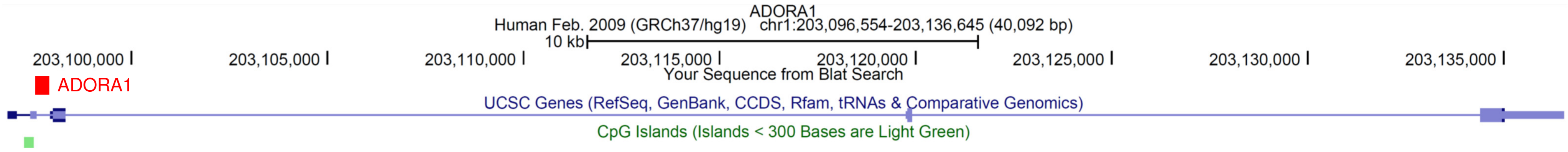

***ADORA1*:** DNA methylation of 7 CpGs in *ADORA1* (Adenosine A1 Receptor) was analyzed (chr1:203.097.582-203.097.833).

##### 3.1.6 *TSPAN9*

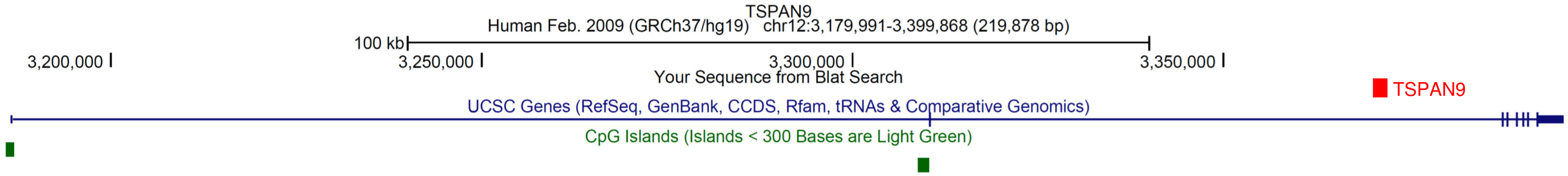

***TSPAN9:*** DNA methylation of 6 CpGs in *TSPAN9* (Tetraspanin 9) were analyzed (chr12:3.370.787-3.371.092).

##### 3.1.7 *RPS6KA2*

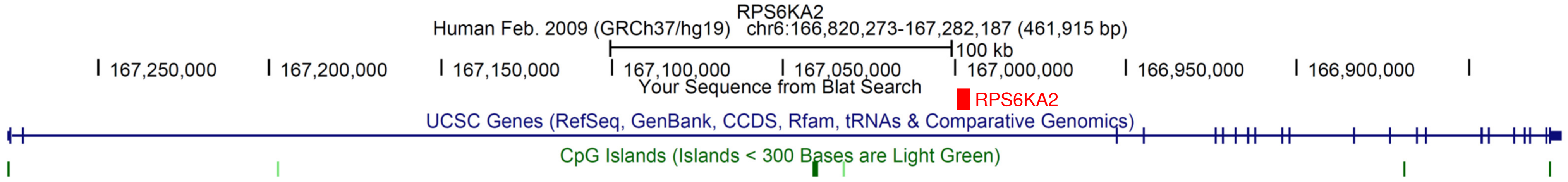

***RPS6KA2:*** DNA Methylation of 8 CpGs in *RPS6KA2* (Ribosomal Protein S6 Kinase A2) was analyzed (chr6:166.996.725-166.997.045).

##### 3.1.8 DMR-1

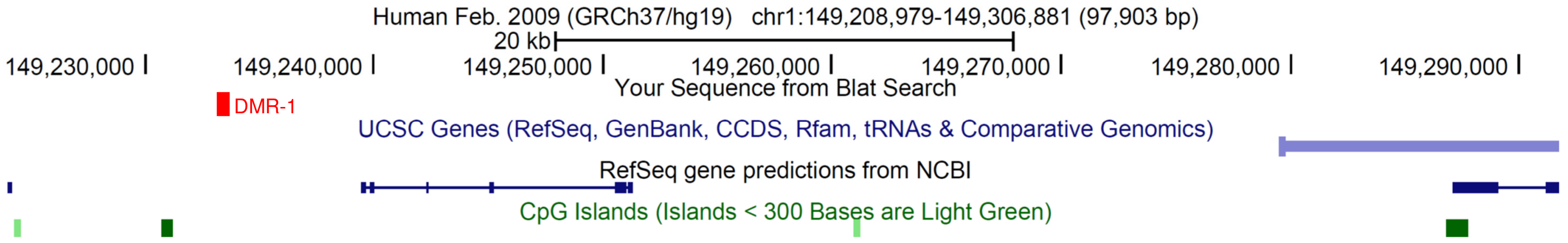

**DMR-1:** DNA methylation of 14 CpGs was analyzed according to the WGBS results. The differentially methylated region (DMR) is about 9 kbp downstream of *RNVU1-18* (RNA. Variant U1 Small Nuclear 18). 6 kbp upstream of *LOC105369140* (NBPF Member 6 Pseudogene) and ~50 kbp upstream of Long Intergenic Non-Protein Coding RNAs (*LOC388692* and *LOC644634*). *RNVU1-18* is affiliated with the snRNA class. *LOC105369140* is a pseudogene. Location of DMR-1: chr1:149.233.322-149.233.624. CpG sites which are influenced by SNPs were not analyzed (rs2319160 and rs2319163).

#### 3.2 S2 Fig. Correlations between symptom change and DNA methylation change in the PTSD cohort

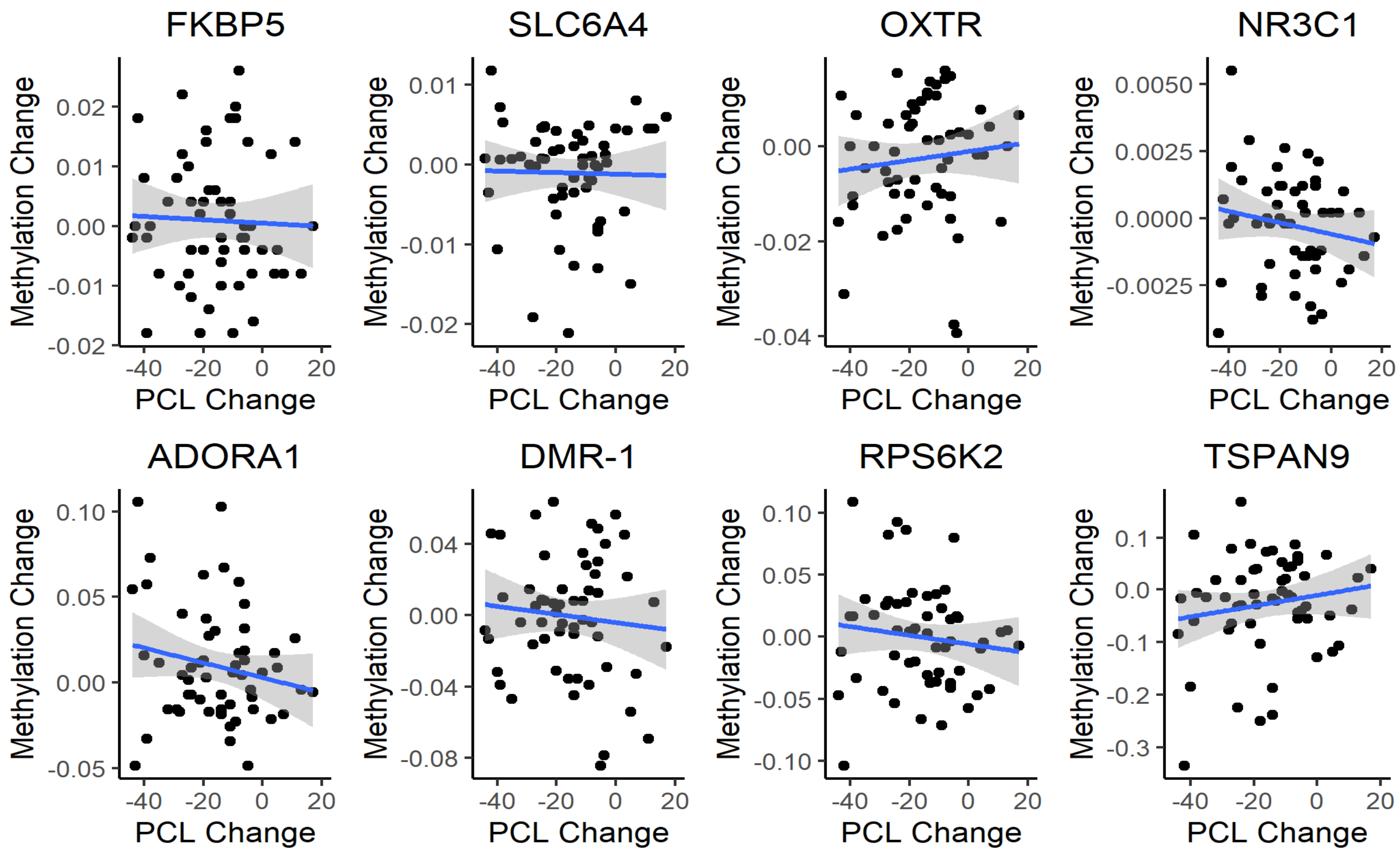

#### 3.3 S3 Fig. Mean DNA Methylation of the PTSD cohort

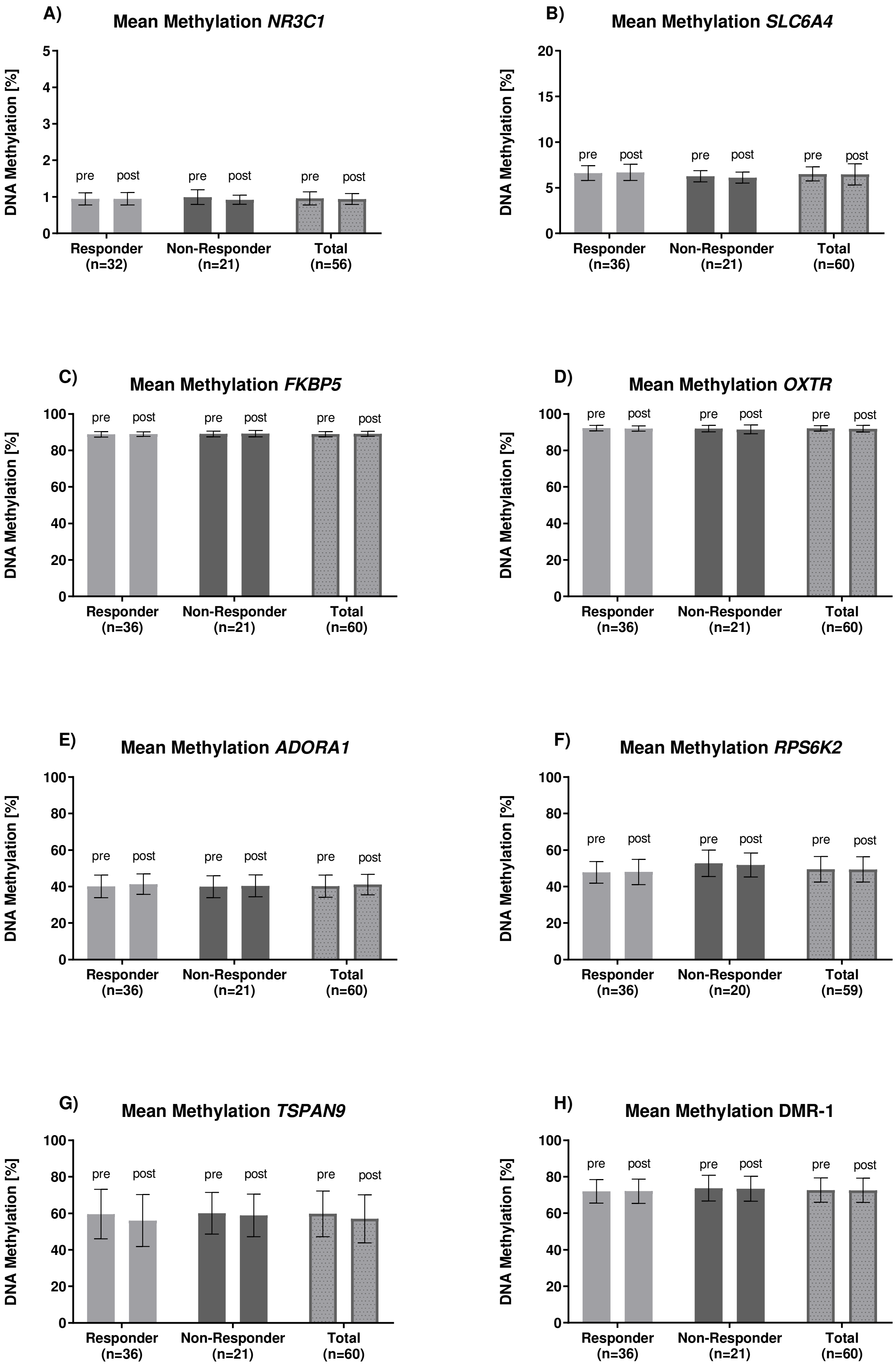

#### 3.4 S4 Fig. Principal component analysis

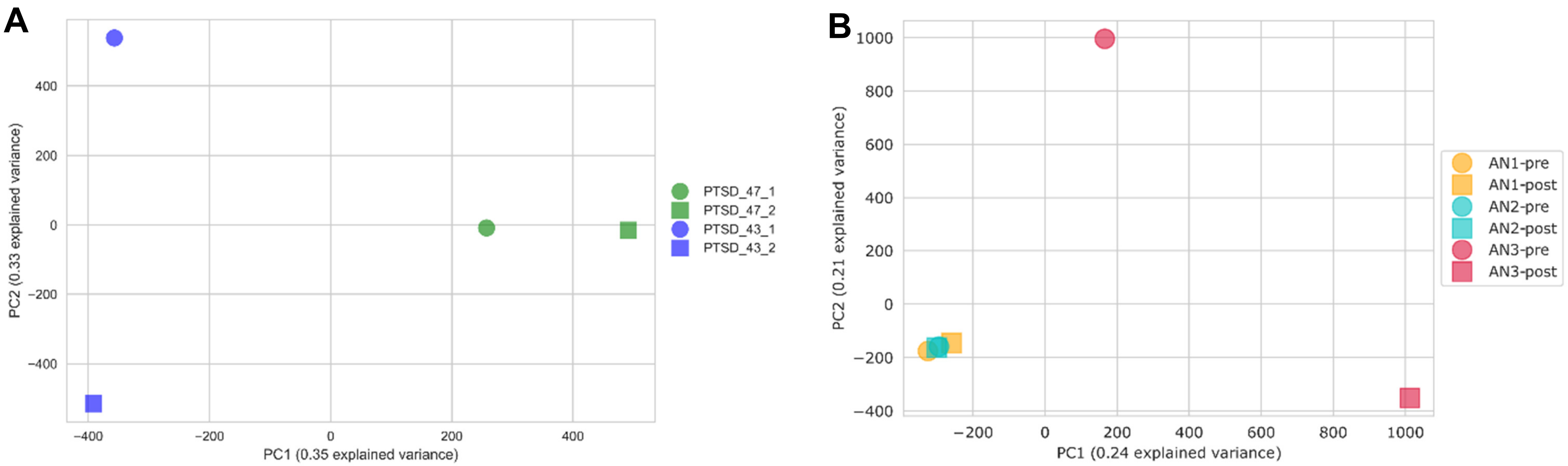

**S4 Fig. Principal component analysis.** (PCA): A) PCA of the four PTSD datasets (2.8 million CpGs). B) PCA of the six AN datasets (23 million CpGs). Only CpG loci with minimum coverage of 10 reads in all samples and minimum mapping quality of 30 are considered.

##### 3.5 S5 Fig. DNA methylation differences of the AN cohort

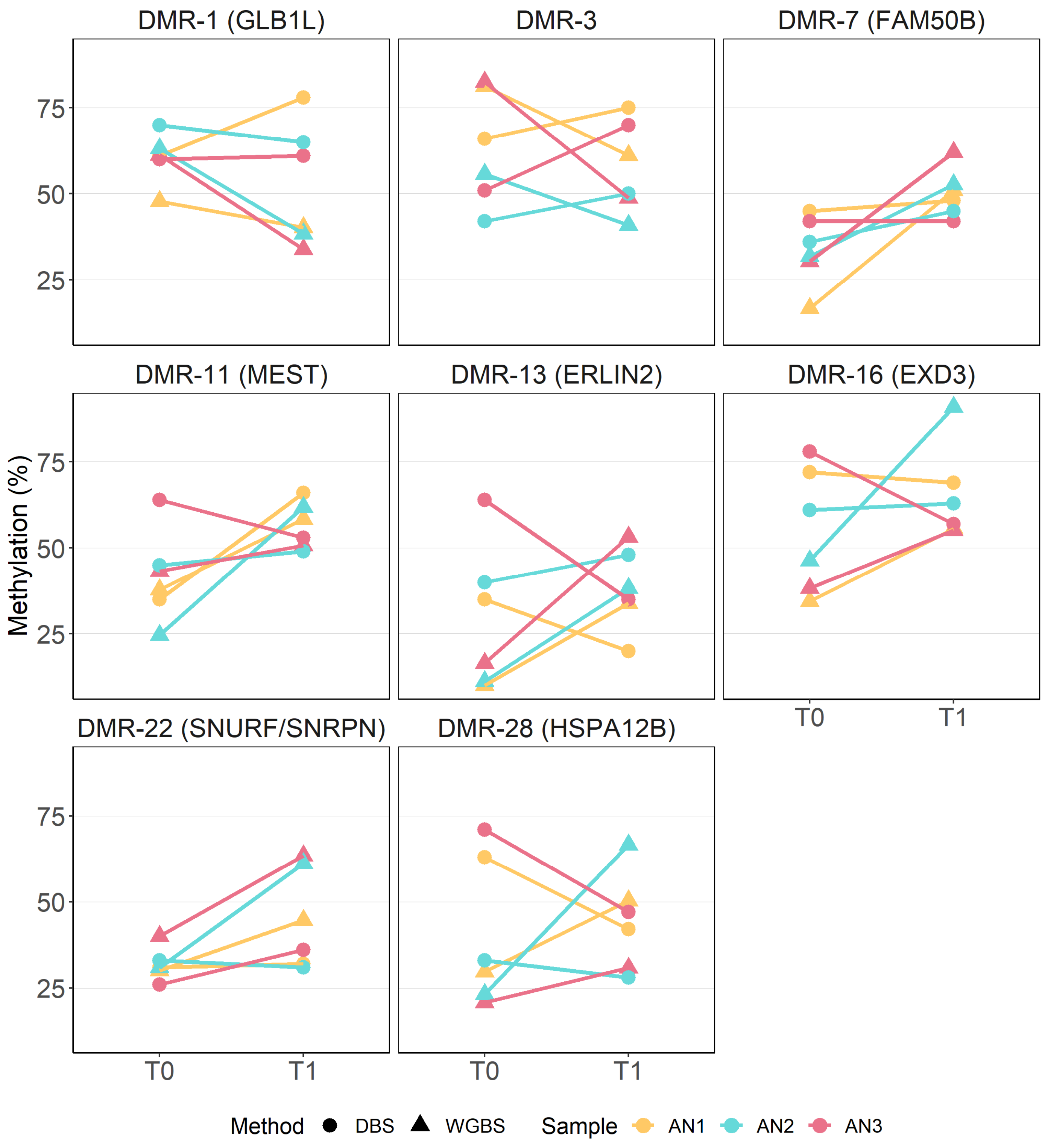

**S5 Fig. Analysis of candidate AN_DMRs by WGBS and DBS.** T0, before intervention; T1, after intervention.
